# Extreme weather events in the UK and resulting public health outcomes

**DOI:** 10.1101/2024.11.25.24317884

**Authors:** Natalie Dickinson, Llinos Haf Spencer, Shuhua Yang, Caroline Miller, Andrew Hursthouse, Mary Lynch

## Abstract

**Objectives:** Extreme Weather Events (EWEs) are increasingly frequent in the UK and can lead to adverse physical and mental health outcomes which may result in additional pressure on the NHS. The objective of this study is therefore to investigate the health impacts of EWEs on the population in the UK.

**Method:** A systematic review of the evidence was conducted. Seven databases were searched for studies related to the public health outcomes of EWEs.

**Results:** 48 papers met the criteria for inclusion (22 flood, 25 extreme temperature, 1 wind). Flooding is most associated with mental health concerns and associated social effects, whilst temperature extremes impact more on physical health.

**Conclusions:** Whilst there is a great deal of research on the physical and mental health impacts of EWEs in the UK, there is limited evidence on social health impacts, and little consideration of the economic costs of holistic health outcomes. Building resilience against the health impacts of EWEs is essential. Future studies should consider incorporating cost-benefit analyses (CBA) to investigate the economic costs of EWEs on populations and health systems in the UK.

## Introduction

Extreme Weather Events (EWEs), including heatwaves, coldwaves, floods, storms and wildfires, are occurring with increased frequency worldwide, primarily driven by climate change (Walsh et al. 2020). The Intergovernmental Panel on Climate Change (IPCC) state that human-induced climate change has caused greater frequency and intensity of climate extremes globally, which has led to adverse impact on human health (Calvin et al., 2023). The impacts of EWEs in the United Kingdom (UK) are also becoming more pronounced; the UK has witnessed a notable increase in the frequency of severe weather incidents, such as heatwaves, floods and storms (Met Office 2022), with further changes predicted. The latest UK Climate Projections 2018 indicate a higher likelihood of warmer, wetter winters, hotter, drier summers, increased wildfires, more extreme weather events, and rising sea levels (Met Office 2022). Rising temperatures, leading to extreme heat in homes and other buildings, poses an increased risk to human health, wellbeing and productivity, and has been identified as one of the priority risk areas by the UK Climate Change Risk Assessment (UK Government, 2022).

Though a systematic review of EWEs across Europe was recently published (Weilnhammer et al. 2021), given the increase in EWEs in the UK, alongside the public health challenges of the ageing population, the healthcare crisis and socioeconomic deprivation, there is a need to consider the distribution of health implications for the population of the UK specifically, as well as the economic cost of EWEs for the NHS.

### Aim

To investigate the impacts of EWEs on the UK population, public health and wellbeing outcomes. The protocol for this systematic review is on the Prospero database: https://www.crd.york.ac.uk/prospero/display_record.php?RecordID=357688

## Method

Systematic review methods were utilised. The PICO and eligibility criteria are presented in Table 1 (Schardt et al. 2007). Evidence is presented below and the search strategies for this are presented in Supplementary File 1.

**Table 1:** Population, Intervention, Context/Comparator and Outcome (PICO) framework.

| Population | Intervention or exposure | Context/Comparator | Outcome |
| --- | --- | --- | --- |
| Population of the UK affected by EWE | EWEs (defined as heat waves, cold waves, floods, storms, wildfires) | Routine weather events: Normal weather patterns such as average rain, | Tangible evidence relating to the impact of EWEs in the UK |
|  | and windstorms) | sunlight, wind, air pressure | on public health and wellbeing outcomes. |

Databases: Cochrane Library (including the Cochrane Central Register of Controlled Trials (CENTRAL), CINAHL, ASSIA, PsycINFO, PubMed incorporating MEDLINE, Web of Science, and the Database of Abstracts of Reviews of Effects (DARE). Grey literature, including local government reports were also included to limit publication bias, and to ensure that all relevant literature was identified.

Date-range: November 1992 and November 2023.

Data management and Screening: Rayyan reference management software was used to store and manage citations (Rayyan, 2024). Duplicates were removed in the EndNOTE and Rayyan. Citations were screened on title and abstract by four members of the review team (ND, LHS, CM, SY and ML). Full-text articles were retrieved and further assessed for inclusion.

### Inclusion criteria

Papers relating to EWEs’ impact on UK population public health outcomes written in the English language from 1992 to 2023.

### Exclusion criteria

Papers not related to EWEs’ influence on UK population public health and wellbeing outcomes. Any queries regarding inclusion/exclusion were resolved by discussion between members of the review team.

Data extraction: The data was extracted using a pre-defined data extraction tool. The data extraction tables can be found in Supplementary File 2-4. Quality appraisal was conducted by members of the review team using relevant JBI critical appraisal tools e.g. JBI cohort studies checklist (Joanna Briggs Institute 2017). Studies were identified as high, medium and low quality.

## Results

Forty-eight (48) papers met the inclusion criteria for this systematic review; 22 focussed on flood; 25 focussed on temperature and one focussed on wind. Figure 1 shows a PRISMA flow chart (Page et al. 2021). A summary of the included studies can be seen in Table 2.

**Table 2:** Study characteristics of included extreme weather event studies.

| # | Type of weather event | Author | Date | Country | Quality rating |
| --- | --- | --- | --- | --- | --- |
| 1. | Flooding | Carroll et al. | 2010 | England | Moderate |
| 2. | Flooding | Euripidou and Murray | 2004 | England | Low |
| 3. | Flooding | Fewtrell and Kay | 2008 | England | High |
| 4. | Flooding | Fewtrell et al. | 2011 | England | High |
| 5. | Flooding | Findlater et al. | 2023 | England | High |
| 6. | Flooding | Fothergill et al. | 2021 | England | Moderate |
| 7. | Flooding | French et al. | 2019 | England | High |
| 8. | Flooding | Graham et al. | 2019 | England | High |
| 9. | Flooding | Hunter | 2003 | England and Wales | Low |
| 10. | Flooding | Lamond et al. | 2015 | England | High |
| 11. | Flooding | Mason et al. | 2010 | England | High |
| 12. | Flooding | Medd et al. | 2014 | England | Moderate |
| 13. | Flooding | Mehring et al. | 2023 | England | Moderate |
| 14. | Flooding | Milojevic et al. | 2011 | England and Wales | Moderate |
| 15. | Flooding | Milojevic et al. | 2017 | England | Moderate |
| 16. | Flooding | Mulchandani et al. | 2020 | England | High |
| 17. | Flooding | Munro et al. | 2017 | England | High |
| 18. | Flooding | Paranjothy et al. | 2011 | England | High |
| 19. | Flooding | Reacher et al. | 2004 | England | Moderate |
| 20. | Flooding | Robin et al. | 2020 | England | High |
| 21. | Flooding | Tapsell et al. | 2002 | England | High |
| 22. | Flooding | Waite et al. | 2017 | England | High |
| 23. | Heat | Alahmad et al. | 2023 | Worldwide including UK | High |
| 24. | Heat | Arbuthnott and Hajat | 2017 | UK | Moderate |
| 25. | Heat | Berger et al. | 2023 | UK | High |
| 26. | Heat | Bryan et al. | 2020 | Scotland, England and Wales, UK | Moderate |
| 27. | Heat | Cruz et al. | 2020 | UK | Moderate |
| 28. | Heat | Curtis et al. | 2017 | England | Low |
| 29. | Heat | Finlay et al. | 2012 | England | Moderate |
| 30. | Heat | Green et al. | 2016 | England | High |
| 31. | Heat | Hajat et al. | 2002 | England | High |
| 32. | Heat | Heaviside et al. | 2016 | England | High |
| 33. | Heat | Johnson et al. | 2005 | England and Wales | Moderate |
| 34. | Heat | Kovats et al. | 2004 | England | High |
| 35. | Heat | Leonardi et al. | 2006 | England | High |
| 36. | Heat | Oven et al. | 2012 | England | Low |
| 37. | Heat | Page et al. | 2007 | England and Wales | High |
| 38. | Heat | Page et al. | 2012 | UK | High |
| 39. | Heat | Rendell et al. | 2020 | England | High |
| 40. | Heat | Rizmie et al. | 2022 | England | High |
| 41. | Heat | Rooney et al. | 1998 | England and Wales | High |
| 42. | Heat | Sahani et al. | 2022 | England and Scotland | High |
| 43. | Heat | Smith et al. | 2016 | England | High |
| 44. | Heat | Smith et al. | 2016 | England | High |
| 45. | Heat | Thompson et al. | 2022 | England | High |
| 46. | Heat | Wan et al. | 2022 | Scotland | High |
| 47. | Heat | Zhang et al. | 2023 | UK | High |
| 48. | Wind | Goldman et al. | 2014 | OECD countries including UK | Moderate |

Note: OECD is an acronym for Organisation for Economic Co-operation and Development

**Figure 1:** PRISMA study selection flowchart (Page et al., 2021)

The health economists in this authorship team (ML and LHS) were interested in the cost elements of EWEs outcomes. Although the search strategy included for health economic information whilst data extractions were conducted, only one of the papers included a health economic aspect (Disability adjusted life years, DALY’s), but no cost to the NHS specific papers (Fewtrell and Kay, 2008).

Through narrative analysis, three themes were evident to describe the resulting health consequences of EWEs: physical health impacts; mental health impacts and social health impacts. Findings from the studies reviewed will be discussed in the context of these themes in the following section.

## Discussion

### Physical health impacts

#### Temperature Extremes

Heatwaves are the most frequently discussed EWE to impact on physical health. Long periods of heat are known to cause heat strokes and contribute to an increase in other cardio-respiratory health conditions and emergency hospital admissions (Alahmad et al., 2023). A number of quantitative studies have demonstrated the link between excess mortality and extreme heat (Rooney et al., 1998; Johnson et al., 2005; Arbuthnott & Hajat, 2017; Alahmad et al., 2023)., and between hospital admissions and extreme heat (Kovats et al., 2004; Rizmie et al., 2022).

Respiratory illness is the most commonly reported cause of mortality during heatwaves (Rooney et al. 1998; Kovats et al. 2004; Arbuthnott and Hajat 2017). A study in Greater London found increases in emergency admissions for respiratory and renal disease were related to heat, particularly among under 5s, and those over 75 (for respiratory disease) (Kovats et al., 2004). An association between heat and daily mortality was also observed during the 2003 heatwave in England and Wales, marked by a significant short-term rise in deaths. This was particularly notable in London, where mortality among those over 75 increased by 59%. Elevated levels of ozone and particulate matter were also recorded during the heatwave. (Johnson et al., 2005). Furthermore, a study of the 1995 heatwave in England and Wales indicated that up to 62% (London) and 38% (rest of England and Wales) of excess deaths may be attributable to concomitant increases in air pollution (Rooney et al. 1998). Increased heat vulnerability has been observed in areas with high population density, significant urbanization, low coverage of green space, and elevated levels of fine particulate matter (Zhang et al., 2023). Urban heat islands (UHIs) have been found to experience a higher heat-related mortality (up to 50%) more than surrounding areas (Heaviside et al., 2016). The UHI effect is compounded by the coinciding increase in air pollution, further impacting public health. A typical heatwave in 2080 (under a medium emissions scenario) could result in a mortality rate approximately three times higher than that of 2003, when accounting for factors such as population changes, population distribution, and the urban heat island effect, assuming no future changes in heat adaptation (Heaviside et al., 2016).

The links between heat and other health outcomes are less consistent (Arbuthnott and Hajat, 2017). Alahmad *et al*. (2023) observed an increase in the risk for cardiovascular disease (CVD) mortality associated with heat (Alahmad et al. 2023). This study also investigated cold extremes, finding CVD mortality to be four times greater during the cold extremes than hot extremes. A further study of temperature extremes found that a greater number of hospitalisations were correlated with extreme cold than with extreme heat (Rizmie et al., 2022). In contrast to earlier studies, Rizmie *et al*. (2022) found respiratory diseases contributed less to the increased admissions (9.2%) than other conditions; metabolic diseases had the largest effect magnitude (25.8%), followed by infectious diseases (21.1%) at temperatures >30⍰C (Rizmie et al., 2022). Research in Scotland found that mortality risk began to rise when temperatures exceeded 14.5°C, suggesting that heat-related health impacts can occur even at relatively low temperatures in cooler climates (Wan et al. 2022). The authors highlight that in places such as Scotland, where cold extremes are more commonly experienced, heatwaves remain an invisible threat, perhaps highlighted by the absence of a heatwave plan in the devolved nations.

Excess deaths were not observed in England during the 2013 heatwave, when compared to similar sustained heatwaves in 2003 and 2006 (Green et al., 2016), whilst the 1976 heatwave (Hajat et al. 2002) and the 2020 heatwaves (Thompson et al., 2022) were found to be associated with significantly higher numbers of deaths compared to other hot periods. The reasons for this remain unclear, indicating the presence of unknown factors. However, there was no evidence to suggest that the heatwaves increased the proportion of deaths occurring in different settings or that the COVID-19 pandemic altered the primary underlying causes of death during a heatwave. (Thompson et al., 2022).

The demographics of the population experiencing heatwaves influences the health impacts. A review of the UK heatwave literature found the over 65 age group, people with neurological and psychiatric diagnoses, and those living in Greater London, the Southeast and Eastern regions to be more vulnerable to heat (Arbuthnott & Hajat, 2017). These findings are further corroborated by a more recent study, which also discovered that admissions during extreme temperatures were more prevalent among older individuals and, additionally, those from socioeconomically disadvantaged backgrounds. (Rizmie et al. 2022). In a study of NHS 24 calls, which indicated infants and over 65s were more affected by heat, Leonardi *et al*. (2006) highlight that the morbidity of the elderly in relation to heat is underrepresented in hospital records, as this demographic may be less able to perceive ambient temperature, and be less inclined to seek healthcare services (Leonardi et al. 2006). This suggests there may be a need for social heath support and pre-emptive intervention for older adults during heatwaves.

One study examined UV exposure and found that its health implications are complex and highly influenced by behaviours and sociodemographic factors, such as skin colour. Public health recommendations could be enhanced by considering both temperature and UV exposure and their effects on behaviour (Rendell et al. 2020). Drought, an effect of prolonged dry weather, can result in reduction in water quantity and quality, thereby negatively impacting health and well-being through compromised hygiene and sanitation, food security, and air quality (Bryan et al., 2020). Wildfires, a less common consequence of extreme heat, can severely impact human health through inhalation of wood smoke, which impacts particularly on vulnerable populations (respiratory, cardiovascular, ophthalmic, psychiatric problems and burns (Finlay et al., 2012).

#### Windstorms

Only one moderate quality review investigated the direct health outcomes occurring during a powerful windstorm (Goldman et al., 2014). Human health can be significantly impacted by storms; direct effects occur during the storm’s impact phase, resulting in deaths and injuries from the force of the wind. Key dangers include individuals becoming airborne, being struck by flying debris or falling trees, and road traffic accidents. Furthermore, the exacerbation of chronic illnesses due to limited access to medical care or medication can also adversely affect health outcomes (Goldman et al., 2014).

#### Flooding

Flooding in the UK is linked with contaminated water, leading to gastrointestinal illness supplies (Hunter 2003). Cryptosporidium has been found to be the most commonly reported pathogen in public water supply, with Campylobacter being the commonest cause of outbreaks in private water supplies (Hunter 2003). More recent evidence suggests that up to 3% of the flooded population will have a gastrointestinal illness after swallowing cumulative amounts of floodwater during the clean- up process. The use of gloves could help to reduce contact with biological contaminants, though disposable facemasks were suggested to be more effective (Fewtrell et al., 2011). In addition, findings of a UK study reveal that flooding of an urban river with input from a wastewater treatment works above the flooding point was likely to result in greater gastrointestinal effects than flooding caused by run-off from a more microbiologically pristine area (Fewtrell & Kay, 2008).

A literature review in 2004 highlighted three flood related chemical incidents in England following flooding, with health effects including sore throat, nausea, and stomach pains along with skin irritation. The review led to the development of a checklist for public health response and investigation for extreme flooding, which included chemical contamination identification events (Euripidou & Murray, 2004). No recent research was found linking chemical contamination with flooding.

An investigation of the long-term effects of flooding on mortality in England and Wales found an unexpected deficit of deaths in the year after flooding (post-/pre-flood ratio of 0.90, 95% CI 0.82, 1.00). No significant variation was found by age, sex, population density or deprivation, and the authors suggest displacement may be the reason for the ‘deficit of deaths’ within the same areas pre and post flood (Milojevic et al., 2011).

### Mental health impacts

#### Extreme Temperatures

Though heatwaves can have positive impacts on mental health, they can also have negative consequences (Bryan et al., 2020). A study found that when temperatures exceeded 18°C, each 1°C rise in the average temperature was linked to a 4% increase in suicide and a 5% rise in violent suicides. During the 1995 heatwave, suicide rates surged by 47%, while no significant change was observed during the 2003 heatwave (Page et al., 2007). In addition, research has shown an overall increase in risk of death of 5% per 1°C increase in temperature (95% CI 2.0–7.8) in people with psychosis, dementia, and substance misuse (Page et al., 2012). The greatest mortality risk was observed in younger patients and those with a diagnosis of substance misuse, with the authors noting that heatwave public health strategies should be targeted towards these groups. Whilst none of the selected studies referred to mental health impacts of cold temperatures, this is likely due to the selection criteria for this review; the effect of cold weather on mental wellbeing in the UK is extensively studied, but in relation to sustained cold temperatures rather than ‘extreme cold’ or ‘coldwaves’.

#### Flooding

Mental health impact is the most significant effect of flooding (Euripidou and Murray 2004). This negative effect has been demonstrated quantitatively in a number of studies (Reacher et al., 2004; Fewtrell and Kay, 2008; Mulchandani et al., 2020; Graham et al., 2019). Graham *et al*. (2019) utilised a national mental health survey to compare incidence of common mental disorder (CMD) and sociodemographic characteristics of those who had experienced flood-related damage to the home within the past six months, with those who had not. A multiple regression model, controlling for socioeconomic factors and health status, found an increase of 50% in the odds of CMD (OR 1.5, 95%CI 1.08; 2.07) in those who experienced flood damage.

The effect of flooding has been shown to have lasting effects. One year after flooding, prevalence of psychological morbidity in flooded participants has been observed (depression 20.1%, anxiety 28.3% and PTSD 36.2%) (Waite et al., 2017). When compared with unaffected participants, adjusted odds ratios showed psychological morbidity to be 6-7 times higher in flooded than unaffected participants (aOR (95% CI): depression 5.91 (3.17-10.99); anxiety 6.50 (3.77–11.24); PTSD 7.19 (4.33-11.93), and 1-2 time higher in participants disrupted by the floods compared to those unaffected (aOR (95% CI): probably depression 1.56 (0.88-2.76); probable anxiety 1.61 (0.94–2.77); probable PTSD 2.06 (1.27- 3.35) (Waite et al., 2017). Similarly, it has been found that even three years after flooding, 7.9% of flooded respondents had probable depression, 11.7% had probable anxiety and 17.5% had probable PTSD, with higher prevalence in the flooded group compared with the unaffected group (Mulchandani et al. 2020). After adjustment for potential confounders, probable mental health outcomes were higher in the flooded group compared to the unaffected group, significantly for probable depression (aOR (95% CI) 8.48 (1.04–68.97) and PTSD (aOR (95% CI) 7.74 (2.24–26.79)) (Mulchandani et al., 2020). This is further supported by another study which used Health Related Quality of Life (HRQoL) scores (Robin et al., 2020). Median HRQoL scores were lower in the flooded and disrupted groups compared with unaffected respondents at both two and three years post-flooding. After two years, associations between exposure to flooding and experiencing anxiety or depression were observed (aOR (95% CI) 7.7 (4.6–13.5)), persisting, though less apparent, three years post-flooding (aOR (95% CI) 4.3 (2.5–7.7)) (Robin et al., 2020).

The prevalence of mental health conditions appears to increase with the presence and level of flood water in the home (Paranjothy et al., 2011). All measured mental health symptoms were higher (p <0.01) in those who had flood water in their home than those who did not. Those with flood water above floor level had higher odds of mental health conditions (aOR (95% CI) 12.8 (9.3–17.6)) than those with water below floor level (aOR (95% CI) 3.0 (2.0–4.6)). Similarly, Waite *et al*. (2017) observed increased depth of floodwater to be significantly associated with all mental health conditions; depression, for example, ranged from aOR (95% CI) 4.58 (2.38-8.80) for flood depth <30cm, to aOR (95% CI) 8.48 (4.21-17.10) for 30-100cm, and aOR (95% CI) 14.71 (4.45 – 48.62) for flood depth >100cm. Paranjothy et al. (2011) also highlight subsequent disruption to services and power, more often experienced with greater flood depths, to be negatively associated with mental health outcomes.

Displacement from the home has been found to worsen flooding’s impact on mental health. Those who had to relocate for over six months were six times more likely to experience mental health issues than those that did not need to relocate (Lamond et al., 2015). Two further studies support this, finding displacement from home to be significantly associated with higher scores for depression, anxiety and PTSD, and individuals with poor health who had to vacate their home, along with having no previous knowledge of flooding experienced greatest levels of psychological distress (Mason et al., 2010; Munro et al., 2017).

The prevalence of depression was higher among respondents who experienced repeated flooding compared to those affected only once, but this difference was not statistically significant after adjustment. Additionally, there were no differences in anxiety or PTSD levels, and only minimal differences in overall HRQoL between the two groups (French et al., 2019). It could be suggested that experiencing flooding on one occasion allows those affected to build resilience for future similar events, indeed authors have observed that those without prior knowledge of flooding experienced greater distress (Paranjothy et al., 2011). Individuals who experienced a flood reported feeling unprepared and unsure of how to safely deal with flood water and potentially contaminated possessions (Cruz et al., 2020). Depression and PTSD scores were higher in individuals who were displaced and did not receive any warning, compared to those who were warned more than 12 hours before the flooding. Though the difference in anxiety scores was not significant, these findings suggest that receiving advanced warning may offer protection against mental health issues (Munro et al., 2017).

Exploratory research has sought to explore the underlying mechanisms linking flooding to mental health outcomes. It has been observed that it is often not the flooding itself, but the recovery process that can be difficult to deal with: ‘project managing’, ‘fighting’, ‘loss of treasured possessions’, and ‘stripping-out the home’ (Medd et al. 2014). This study underlined the value of understanding and developing the flood recovery process to enhance resilience and preparedness for future floods. It is crucial to provide support and space for flood victims to share their experiences and be heard, which can assist in their recovery and contribute to better flood management strategies. Various authors support the notion that the after-effects of flooding on mental health may be prolonged, as those affected not only need to rebuild their lives after flooding, but also need to build resilience to cope with the continued threat of living in a flooding hot spot (Medd et al. 2014; Fothergill et al. 2021; Mehring et al. 2023). Interventions that empower individuals living with ongoing flood risk are thought to potentially enhance psychological resilience (Fothergill et al. 2021). It has been suggested that prioritising long-term psychological support from both formal and informal sources should be central to strategies addressing the impacts of flooding; flooding is likely to increase demand for primary care, counselling, and voluntary services, while also placing considerable strain on informal support networks (Findlater et al., 2023), therefore targeting vulnerable groups is essential. Additionally, reinstating access to public transport, education, work, and health and social care services as soon as possible may be protective against mental health morbidity (Waite et al. 2017)

### Social health

The literature consistently shows that various socio-economic factors, such as income, age, gender, pre-existing health conditions, and family structure, are linked to increased vulnerability to flooding (Tapsell et al. 2002; Paranjothy et al. 2011; Medd et al. 2014). Women, ethnic minorities, those with lower income levels and older people (aged over 65 years) were found to be more affected (Cruz et al. 2020). The effects of flooding on mental health are not equally distributed; the impacts vary according to gender comorbidities, socioeconomic status and level of damage or disruption experienced during an EWE. Females have been found to score more highly on PTSD, anxiety and depression scores than males, as have those in poor general health (Mason et al., 2010). Mental health conditions were 3-5 times higher among the more socio-economically deprived Yorkshire study area than in Worcestershire, and were significantly more likely in women, unemployed, and people with comorbidities (Paranjothy et al., 2011). The importance of risk factors for common mental health disorders was similarly observed by Graham *et al*. (2019), specifically for females, those living in a deprived area, financial debt, comorbidities, and alcohol abuse (Graham et al., 2019). Findings also suggest that lower income households are less likely to report storm or flood damage to their homes, or may be less likely to have contents insurance, resulting in under- representation in the figures generated by insurance companies (Graham et al., 2019).

Evidence highlights the inadequacy of modelling studies to measure and predict flood damage and recovery, as they cannot account for non-measurable, intangible aspects of flooding, nor for the secondary impacts encountered during the recovery process (Tapsell et al. 2002; Medd et al. 2014); both studies pointed to the meaning of home and neighbourhood, and how that can change throughout the recovery process. Higher odds of mental illness have been associated with a disruption to health and social care, and work/education amongst those living in flood-affected areas (Waite et al., 2017). Instead of experiencing a steady progression toward normalisation, participants in a qualitative study described a pattern of highs and lows influenced by other life events, and placed importance on the quality of their interactions with the agencies involved in the recovery process (Carroll, Balogh, et al., 2010). The wider community is also affected, with front line workers report suffering from overwork, stress, and emotional turmoil, and that support was unavailable (Carroll, Balogh, et al., 2010).

Geographical patterning of deprivation tends to see seaside towns housing the most deprived communities, whilst riverside locations tend to be home to the most affluent in society (Graham et al., 2019). A mapping study revealed that areas most at risk of significant future flooding increases were coastal or situated along major estuaries, rather than those prone to river flooding (Oven et al., 2012), further compounding inequalities. The Scottish population is understudied relative to England, yet people in Scotland may be more vulnerable to temperature extremes due to the already higher mortality rate (inequality, negative health behaviours, deprivation) (Wan et al., 2022).

Temperature extremes are also not experienced equally. Not only has it been suggested that higher nighttime temperatures associated with greater socio-economic deprivation may contribute to excess mortality (Rooney et al., 1998), research has found infant sleep to be compromised during hot weather, with the authors highlighting that even mild sleep deprivation can negatively impact concentration and learning ability (Berger et al. 2023). Considering those from lower socio-economic groups are again likely to be the most disadvantaged, this has potential to further widen the inequality gap.

Future research should consider housing conditions, socioeconomic conditions and segregating mortality type (by disease/age/sociodemographic factors) to calculate risk for vulnerable populations regarding extreme heat (Sahani et al., 2022). Preventative approaches to build resilience within communities are recommended (Cruz et al., 2020).

## Conclusion

Though the evidence of EWEs impact on health is overwhelming, little progress has been made in recent years to mitigate the impacts. The needs are clear: joined -up, multi-agency research and future planning is essential to allow the UK’s health service to meet the needs of the population. Integrated Health and Social Care Partnerships which focus on anticipatory and preventative care (UK Government 2022b; Scottish Government 2024) will no doubt benefit holistic health. There is a need for climate-related health planning to consider the impact on the ageing populations, given those over 65 are more affected by EWEs (Kovats and Brisley 2021). This suggests a need for closer proactive monitoring of older people during EWEs, and may provide an opportunity for the use of telehealth methods, in line with current strategy (Scottish Government 2021).

Further research on preparedness for EWEs is needed, including consideration of how to record and compile information about EWEs and develop risk assessments, assess vulnerabilities and adaption over time (Stott et al. 2016; Clarke et al. 2021). Syndromic surveillance is becoming popular as a method of tracking impacts of EWEs (Curtis et al., 2017). There is potential for complementing experiential learning with targeted mitigation support to build resilience, especially in communities likely to experience further flooding (Lamond et al. 2015). Recognising that interventions which rely on behavioural change are unlikely to be equally adopted, thereby exacerbating inequalities, there is a need to plan for supporting communities, particularly low-income families, to prepare for EWEs (Kovats & Brisley, 2021; Marmot, 2020).

Preparedness should include response strategies from health and social care providers as well as emergency response strategies (Curtis et al., 2017). To meaningfully plan to support those affected by EWEs, there is a need to build NHS costs and psycho-social impacts into future cost-benefit analyses in prospective EWE studies examining health impacts and outcomes. There is scope to support the development of policy and interventions for building resilience at a community and societal level, while taking account of health system needs and efficiencies required during and post EWE’s.

### Strengths

- Included evidence on the health outcomes of EWEs from the past 30 years.
- Health outcomes following EWEs presented from a UK perspective.

### Limitations

⍰ A limited number of extreme cold studies were included in this review. This might be due to the limitations of the search strategy, and ‘extreme cold’ or ‘cold wave’ not featuring commonly in the literature.
⍰ There was a dearth of included studies from Wales, Scotland and Northern Ireland.

### Recommendations

Future research exploring the impacts of EWEs should include Cost Benefit Analysis (CBA).

Support the integration of health and social care services to provide holistic preventative action against EWE induced health impacts.

Develop strategies to build resilience among individuals, communities and health and care systems for future EWE’s.

Closer monitoring of vulnerable groups.

## Data Availability

All data produced in the present study are available upon reasonable request to the authors.

## Acknowledgements

The authors would like to thank Margo Stewart, Academic Librarian (Health & Life Sciences), Associate Fellow, Higher Education Academy, Library, University of the West of Scotland for providing support to the review team in terms of the search strategy and assisting with the initial searches for this systematic review. The Universities of the West of Scotland, University of South Wales, and the Royal College of Surgeons Ireland supported the researchers time on this systematic review.

## Author Contribution

Conceptualisation: ND and ML; Methodology: ND, LHS, SY and ML. Writing-drafting, reviewing and editing: ND, LHS, SY and ML.

## Disclaimer

The views expressed in this publication are those of the authors, not necessarily The University of the West of Scotland.

