## Supplementary File 1 for "Extreme weather events in the UK and resulting public health outcomes"

### Supplementary file 1 - Search strategy – Extreme Weather Events

**The search strategy for the Extreme Weather Events Systematic Review is presented below.**

“Infant” OR “Child” OR “Children” OR “Boy” OR “Girl” OR “Teenager” OR “Adolescent” OR “Young person” OR “Adult” OR “Female” OR “Male” OR “Transgender” OR “Older person” OR “Person” OR “People” OR “Citizens” OR “Man” OR “Women” OR “Population” OR “Communities” OR “Human”

AND

“Extreme Weather Events” OR “Meteorological conditions” OR “Climate Change” OR “Temperature” OR “Global Warming” OR “Humidity” OR “Greenhouse Effect” OR “Extreme Heat” OR “Natural Disasters” OR “Droughts” OR “Floods” OR “Tornadoes” OR “Cyclonic Storms” OR “Tidal Waves” OR “Tsunamis” OR “Wildfires” OR “Landslides” OR “Extreme Cold Weather” OR “Snow” OR “Rain” OR “Extreme Hot Weather” OR “Sea Level Rise” OR “Adverse weather” OR “Heat stress” OR “Climate events” OR “Contamination” OR “Environmental toxin exposure”

AND

“Morbidity” OR “Mortality” OR “Birth” OR “New conditions” OR “Diverse food-borne infectious diseases” OR “Worsening/improving conditions” OR “Water-borne infectious diseases” OR “Chronic disease” OR “Socio-Economic Status” OR “Injuries” OR “Displacement” OR “Mental health” OR “Suicide” OR “Harm” OR “Drowning” OR “Poverty” OR “Malnutrition” OR “Starvation” OR “Health” OR “Social isolation”

AND

“UK” OR “United Kingdom” OR “Great Britain” OR “Britain” OR “British Isles” OR “Scotland” OR “England” OR “Wales” OR “Northern Ireland”

**END**
