## Supplementary File 2 for "Extreme weather events in the UK and resulting public health outcomes"

### Supplementary file 2 - Evidence table for Extreme Weather Events (EXE) Systematic review (SR) included papers: Flooding

| **#** | **Citation (Country)**  **Aim** | **Intervention details** | **Study characteristics, health economics methods and quality appraisal** | **Outcome and costs measured** | **Main Findings** |
| --- | --- | --- | --- | --- | --- |
| 1. | Carroll, B., Balogh, R., Morbey, H., & Araoz, G. (2010). Health and social impacts of a flood disaster: responding to needs and implications for practice. Disasters, 34(4), 1045–1063. <https://doi.org/10.1111/j.1467-7717.2010.01182.x>  (Carroll et al., 2010).  **Country:** England, UK  **Aim:** To identify, from  people whose homes had been flooded in Carlisle, Engalnd, the health and social impacts through their perceptions and reported behaviour before, during and after the floods. | **Intervention:** No intervention, this was a qualitative study.  **Dates of data collection:** 2006/2007  **Intervention recipients and sample size:** Five focus group and six  individual interviews were conducted, comprising 14 men and 26 women between the ages of 30 and 70.  **Setting:** Carlisle, North of England, UK.  **Delivery mode (e.g., remotely online, in person):** No intervention was delivered.  **Intervention deliverers:** No intervention was delivered.  **Timing and duration:** All the interviews took place between 10 and 13 months after the floods.  **Intervention description:** No intervention, but people gave their views of living through a flood in the North of England in 2005. | **Study type:** Qualitative study.  **Length of follow-up:** No follow-up.  **Type of economic evaluation/cost analysis:** No economic evaluation.  **Perspective of analysis:** Perspectives of the people who had been flooded in Carlisle, England.  **Currency and cost year:** No costs presented.  **Discounting:** No.  **Sensitivity analysis:** No.  **Quality appraisal:** High | **Outcome/s of interest:** Perceptions of the flooding in Carlisle, England in 2005.  **Types of costs measured:** No costs measured, but human costs described. | **Main finding:** The findings are presented in five sections covering flood risk awareness, water contamination issues, physical health, mental health, and impact on frontline support workers. The discussion focuses on the implications of the findings for policy and practice vis-à-vis psychological health provision, contamination issues, training and support for frontline support workers, matters relating to restoration, and preparation for flooding.  **Additional finding:** Respondents spoke of physical health ailments, psychological stress, water health-and-safety issues related to the floods, and disputes with insurance and construction companies, which they felt had caused and exacerbated psychological health problems.  **Sensitivity analysis results (for economic evaluations):** Not conducted. |
|  | Euripidou,E., Murray,V. (2004). Public health impacts of floods and chemical  Contamination. Journal of Public Health, Vol. 26, No. 4, pp. 376–383. <https://10.1093/pubmed/fdh163>  (Euripidou & Murray, 2004)  **Country:** England  **Aim:** to evaluate the significance of public health risks from chemical contamination following flooding. In addition, reviewed in depth three recent floods related to chemical incidents in England and undertaken a literature review of  flooding events that resulted in chemical contamination in order to develop a checklist to assist public health professionals. | **Intervention:** N/A  **Dates of data collection:** Following fire and floods of 2000 and 2001.  **Intervention recipients and sample size:**  N/A  **Setting:** England (Gloucestershire, Kent, and Yorkshire).  **Delivery mode (e.g., remotely online, in person):** N/A  **Intervention deliverers:** N/A  **Timing and duration:** N/A  **Intervention description:** N/A | **Study type:** Literature review along with the review of three flood related incidents  **Length of follow-up:** N/A  **Type of economic evaluation/cost analysis:** N/A  **Perspective of analysis:** N/A  **Currency and cost year** N/A  **Discounting:** N/A  **Sensitivity analysis: N/A**  **Quality appraisal:** Low | **Outcome/s of interest:** Health impacts and chemical contamination flowing floods.  **Types of costs measured:** N/A | **Main finding:**  Case studies showed  health effects including sore  throat, nausea, stinging faces and stomach pains. Most of the symptoms had resolved 4 weeks after the incident, but some  were still evident 7 months later. Raised anxiety among residents  about chemical contamination was a major issue.  Kent: The health survey showed significant impact  on people’s psychological health and an increase in self-reported symptoms including earache, skin rashes and gastrointestinal  upsets.  **Additional finding:**  A checklist/pro forma  for public health response to and investigation of flooding  events that may result in chemical contamination was  needed.  **Sensitivity analysis results (for economic evaluations):** |
| 2. | Fewtrell, L., D. Kay, D. (2008). An attempt to quantify the health impacts of  flooding in the UK using an urban case study. Public Health, 122(5):443-5 <https://10.1016/j.puhe.2007.09.010>  (Fewtrell & Kay, 2008)  **Country:** England, UK  **Aim:** To quantify the health effects of flooding in the UK to allow comparison between different flooding events. | **Intervention:** This was not an intervention study.  **Dates of data collection:** Within the last 5 years (prior to 2007).  **Intervention recipients and sample size:** 30 households flooded.  **Setting:** England  **Delivery mode (e.g., remotely online, in person):** N/A  **Intervention deliverers:** N/A  **Timing and duration:** N/A  **Intervention description:** N/A | **Study type:** Observational study  **Length of follow-up:** No follow-up period  **Type of economic evaluation/cost analysis:** N/A  **Perspective of analysis:** N/A  **Currency and cost year** N/A  **Discounting:** N/A  **Sensitivity analysis:** N/A  **Quality appraisal:** Moderate | **Outcome/s of interest:** Disability-adjusted life years (DALYs) were used to enable the comparison between different health impacts and different flood events and populations, using two sites subject to pluvial flooding in the Bradford area, UK.  **Types of costs measured:** N/A | **Main finding:** Mental health problems, characterized as psychological distress were found to dominate the calculated health impacts, being considerably greater than the combined physical symptoms in the case study examples.  **Additional finding:** Gastrointestinal effects are likely to be greater as a result of flooding of an urban river with a waste water treatment works input above the flooding point, compared with flooding caused by run-off from a more microbiologically pristine area.  **Sensitivity analysis results (for economic evaluations):** None. |
| 3. | Fewtrell, L., D. Kay, D., Watkins, J., Davies, C., Francis. (2011). The microbiology of urban UK floodwaters and a quantitative microbial risk assessment of flooding and gastrointestinal illness. Journal of Flood Risk Management, 4, 77-87. <https://doi.org/10.1111/j.1753-318X.2011.01092.x>  (Fewtrell et al., 2011)  **Country:** England, UK  **Aim:** To assess the likely cases of gastrointestinal illness resulting from contact with floodwater. | **Intervention:** No intervention.  **Dates of data collection:** Sunday 29 July 2007.  **Intervention recipients and sample size:** Samples of untreated wastewater or wastewater mixed with river water were allowed to stand for  intervals of up to 21 days  **Setting:** Locations in Worcester, Gloucester, Tewkesbury and Upton upon Severn, England, UK.  **Delivery mode (e.g., remotely online, in person):** Water collected by environmental officer.  **Intervention deliverers:** No intervention.  **Timing and duration:** One-off water sampling.  **Intervention description:** One-off water sampling. | **Study type:** Quantitative (simulated quantitative microbial risk assessment).  **Length of follow-up:** No follow-up  **Type of economic evaluation/cost analysis:** No economic analysis.  **Perspective of analysis:** N/A  **Currency and cost year:** N/A  **Discounting**: N/A  **Sensitivity analysis:** N/A  **Quality appraisal:** High | **Outcome/s of interest:**  To identify the pathogen concentrations to which people are exposed during flooding along with the degree of gastrointestinal illness resulting from contact with floodwater.  **Types of costs measured:** N/A | **Main finding:** It was estimated that almost 3% of the flooded population will have a gastrointestinal illness after swallowing cumulative amounts of floodwater during the clean-up process. Sampling during flood  events of water and sediment would strengthen the practical policy evidence base required for assessing the severity and range of health impacts using risk assessment methodology.  **Additional finding:** Gloves may help to reduce such contact, although a disposable facemask is likely to be more effective, if less pleasant to wear.  **Sensitivity analysis results (for economic evaluations):** None. |
| 4. | Findlater, L., Robin, C., Hopgood, K., Waite, T., National Study of Flooding and Health Study Group Waite Thomas Beck Charles Oliver Isabel Amlôt Richard Bone Angie Leonardi Giovanni Rubin Gideon James Kovats Sari Armstrong Ben, Rubin, G., ... & Oliver, I. (2023). Help-seeking following a flooding event: a cross-sectional analysis of adults affected by flooding in England in winter 2013/14. *European Journal of Public Health*, ckad082. [**https://doi.org/10.1093/eurpub/ckad082**](https://doi.org/10.1093/eurpub/ckad082)  (Findlater et al., 2023)  **Country:** England, UK  **Aim:** The aim was to describe the help-seeking behaviour of individuals affected by flooding up to 3 years after exposure, which could lead to an additional demand for health and care services when compared to people unaffected by flooding. | **Intervention**: No intervention.  **Dates of data collection:** Winter 2013/ 14.  **Intervention recipients and sample size:** Participants (Year 1: n=2006; Year 2: n=988; Year 3: n=819).  **Setting: England**  **Delivery mode (e.g., remotely online, in person):** This cross-sectional analysis is based on self-completed questionnaires sent in January 2015—8761 households in postcode areas known to have been affected by flooding between 1 December 2013 and 31 March 2014 as part of the National Study of Flooding and Health.  **Intervention deliverers:** No intervention.  **Timing and duration:** Winter 2013-2014.  **Intervention description:** No intervention. | **Study type:** Cross-sectional.  **Length of follow-up:** No follow-up period. Cross-sectional only.  **Type of economic evaluation/cost analysis:** None (frequency of health seeking behaviour only).  **Perspective of analysis:** N/A  **Currency and cost year:** N/A  **Discounting**: N/A  **Sensitivity analysis:** No.  **Quality appraisal:** High | **Outcome/s of interest:** Help seeking behaviour.  **Types of costs measured:** No costs measured (just frequency of health seeking behaviour). | **Main finding:** This study indicates that there is likely to be an increased demand on primary care, counselling, and voluntary services as well as an impact on informal sources of support, following flooding. This effect can be detected at least 3 years post- flooding or disruption.  **Additional finding:**  This research suggests that addressing the long-term and often unmet need for psychological support, from both formal and informal sources, should be paramount in any strategy to support.  **Sensitivity analysis results (for economic evaluations):** No economic analysis conducted. |
| 5. | Fothergill, L. J., Disney, A. S., & Wilson, E. E. (2021). A qualitative exploration of the psychological impacts of living with the uncertainty of persistent flood risk. *Public Health*, *198*, 141-145. DOI: [**10.1016/j.puhe.2021.07.016**](https://doi.org/10.1016/j.puhe.2021.07.016)  (Fothergill et al., 2021)  **Country: England, UK**  **Aim:** To generate insight into individual experiences of living with persistent flood risk, how it affects psychological well-being, and the forms of support deemed appropriate to mitigate psychological risks. | **Intervention:** No intervention.  **Dates of data collection:** 1999-2000.  **Intervention recipients and sample size:** 40 participants were interviewed.  **Setting:** Nottinghamshire, England.  **Delivery mode (e.g., remotely online, in person):** In Person.  **Intervention deliverers:** No intervention  **Timing and duration:** No intervention  **Intervention description:** No intervention. | **Study type:** Qualitative.  **Length of follow-up:** No follow-up.  **Type of economic evaluation/cost analysis:** No economic analysis  **Perspective of analysis:** N/A  **Currency and cost year:** N/A  **Discounting:** N/A  **Sensitivity analysis:** No.  **Quality appraisal:** Moderate | **Outcome/s of interest:** Perceptions and experiences of those living with flood risk.  **Types of costs measured:** No costs measured. | **Main finding:** Living with the uncertainty of persistent flood risk can have significant psychological impacts.  **Additional finding:** Interventions that facilitate the empowerment of individuals living with persistent flood risk may strengthen psychological resilience.  **Sensitivity analysis results (for economic evaluations):** No economic analysis conducted. |
| 6. | French CE, Waite TD, Armstrong B, Rubin GJ., Beck CR, Oliver, I. (2019). Impact of repeat flooding on mental health and health-related quality of life: a cross-sectional analysis of the English National Study of Flooding and Health. BMJ Open. 2;9(11):e031562. <https://10.1136/bmjopen-2019-031562>  (French et al., 2019)  **Country:** England, UK  **Aim:** To assess the association between flooding/ repeat flooding and: (1) psychological morbidity (anxiety, depression, post-traumatic stress disorder (PTSD)) and (2) health-related quality of life (HRQoL) at 6months post flooding. | **Intervention:** No intervention.  **Dates of data collection:** June 2016.  **Intervention recipients and sample size:**  2500 residents (n = 590 people).  **Setting:** Cumbria, England, UK.  **Delivery mode (e.g., remotely online, in person):** N/A  **Intervention deliverers:** N/A  **Timing and duration:** 6 months post-flooding.  **Intervention description:** N/A | **Study type:** A cross-sectional study (survey).  **Length of follow-up:** Only one time point – 6 months after the floods.  **Type of economic evaluation/cost analysis:** N/A  **Perspective of analysis:** N/A  **Currency and cost year:** N/A  **Discounting:** N/A  **Sensitivity analysis:** N/A  **Quality appraisal:** High | **Outcome/s of interest:**   - Depression - Generalised anxiety - PTSD - General health and well-being   **Types of costs measured:** N/A | **Main finding:** Mental health outcomes were elevated among flooded compared with unaffected participants (adjusted OR for probable depression: 7.77, 95%CI: 1.51 to 40.13; anxiety: 4.16, 95%CI: 1.18 to 14.70; PTSD: 14.41, 95%CI: 3.91 to 53.13). The prevalence of depression was higher among repeat compared with single flooded participants, but this was not significant after adjustment. There was no difference in levels of anxiety or PTSD. Compared with unaffected participants, those flooded had lower EQ-5D-5L  index scores (adjusted coefficient: −0.06, 95%CI: −0.12 to −0.01) and lower self-rated health scores (adjusted coefficient: −6.99, 95%CI: −11.96 to −2.02).  **Additional finding:** There was little difference in HRQoL overall between repeat and single flooded participants.  **Sensitivity analysis results (for economic evaluations):** None. |
| 7. | Graham, H., White, P., Cotton, J., & McManus, S. (2019). Flood- and weather-damaged homes and mental health: An analysis using England’s mental health survey. International Journal of Environmental Research and Public Health, 16(18). https://doi.org/10.3390/ijerph16183256  (Graham et al., 2019)  **Country:** England, UK  **Aim:** To investigate the social patterning of storm and flood damage.  To establish whether recent damage to the home from storms and floods independently predicted Common Mental Disorder (CMD). | **Intervention:** No intervention.  **Dates of data collection:**  2014-2015.  **Population and sample size:**  Analysis of the Adult Psychiatric Morbidity Survey 2014-2015) (n = 7525).  354 (4.5%) of participants answered that they had experienced damage to their home from wind, rain, snow, or flood (only 0.1% displaced).  **Setting:** Nationwide  **Delivery mode (e.g., remotely online, in person):** -No intervention.  **Intervention deliverers:** No intervention.  **Timing and duration:**  Cross section. Survey taken in 2014-15 covers the period of December 2013 – March 2014, when there was extensive flooding in the UK.  **Intervention description:** No intervention | **Study type:** Quantitative. Cross sectional postal survey.  **Length of follow-up:** No follow-up.  **Type of economic evaluation/cost analysis:** No economic analysis.  **Perspective of analysis:** N/A  **Currency and cost year:** N/A  **Discounting:** N/A  **Sensitivity analysis:** N/A  **Quality appraisal:** High | **Outcome/s of interest:**  Common Mental Disorder (CMD) or PTSD. Flood-exposed vs non-exposed adults.  **Types of costs measured:** No costs measured.  **Type of health impacts measured:**  **Physical** –  **Mental** – CMD or PTSD  **Social** –  **Economic** Financial- low income increases risk of CMD. | **Main finding:**  Storm or flood damage significantly associated with CMD (p < 0.01). This association remained after adjusting for socio-demographics, socio-economics, area level.  **Additional finding:**  Confirms importance of known risk factors, for CMD, including being female, aged 16-24, living in a deprived neighbourhood, being in debt, having poorer general health, and harmful patterns of alcohol abuse.  Too small a sample (0.1%) displaced by flooding, so no further analysis on this population.  Advantaged groups more likely to report storm or flood damage to their homes. This differs from most other findings. The authors point out that the 2013-14 floods were predominantly in the affluent southern counties. They also highlight the need to distinguish between tidal flooding and fluvial flooding, as the geographical patterning of deprivation tends to see seaside towns housing the most deprived communities, whilst riverside locations tend to be home to the most affluent.  **Sensitivity analysis results (for economic evaluations):**  **Recommendations:** No economic analysis conducted. |
| 8. | Hunter, P. R. (2003). Climate change and waterborne and vector‐borne disease. *Journal of applied microbiology*, *94*(s1), 37-46. DOI: [**10.1046/j.1365-2672.94.s1.5.x**](https://doi.org/10.1046/j.1365-2672.94.s1.5.x)  (Hunter, 2003)  **Country:** England and Wales, UK  **Aim:** To investigate the burden of waterborne disease in the UK from recorded outbreaks. | **Intervention:** No intervention  **Dates of data collection:** 1991-2000  **Intervention recipients and sample size:** n = 4,112 people who had an infection linked to water.  **Setting:** Worldwide.  **Delivery mode (e.g., remotely online, in person):** Secondary data.  **Intervention deliverers:** No intervention.  **Timing and duration:** No intervention.  **Intervention description:** No intervention. | **Study type:** Review of evidence.  **Length of follow-up:** No follow-up.  **Type of economic evaluation/cost analysis.** No economic analysis.  **Perspective of analysis:** N/A  **Currency and cost year:** N/A  **Discounting:** N/A  **Sensitivity analysis:** No.  **Quality appraisal:** Low | **Outcome/s of interest: I**nfections liked to dirty water:   - Acanthamoeba keratitis - Leptospirosis - Hepatitis E - Malaria - Diarrhoeal disease - Acute respiratory disease   **Types of costs measured**: No costs measured. | **Main finding:** Main finding: The evidence about the burden of waterborne disease in the UK comes predominantly from recording of outbreaks. In the UK there has been some 65 recorded outbreaks of infection linked to water affecting 4112 people during the years 1991–2000. Of these outbreaks, 25 were associated with public water supplies, 16 with private water supplies, 23 with swimming pools and one with recreational contact with surface waters.  **Additional finding:** By far the commonest reported pathogen was Cryptosporidium, although Campylobacter was the commonest cause of outbreaks associated with private water supplies.  **Sensitivity analysis results (for economic evaluations):** No economic analysis conducted. |
| 9. | Lamond, J. E., Joseph, R. D., & Proverbs, D. G. (2015a). An exploration of factors affecting the long term psychological impact and deterioration of mental health in flooded households. Environmental Research, 140, 325–334. <https://doi.org/10.1016/j.envres.2015.04.008>.  (Lamond et al., 2015)  **Country:**  England, UK  **Aim:** To improve understanding of the importance of the recovery experience in mental health outcomes. | **Intervention:** No intervention.  **Dates of data collection:**  2013  **Population and sample size:** UK wide, locations flooded during 2007 floods.  280 responses (household level) (12.1% response rate).  **Setting:** Clusters in the North of England (including Sheffield and Hull) and in the South (including Gloucester and Swindon).  **Delivery mode (e.g., remotely online, in person):** - No intervention.  **Intervention deliverers:** No intervention.  **Timing and duration:**  Cross sectional, 6 years post-flood.  **Intervention description:** No intervention. | **Study type:** Quantitative. Survey of flooded households 6 years post-flooding.  **Length of follow-up:** No follow-up.  **Type of economic evaluation/cost analysis:** No costs measured.  **Perspective of analysis:** N/A  **Currency and cost year:** N/A  **Discounting:** N/A  **Sensitivity analysis:** N/A  **Quality appraisal:** High | **Outcome/s of interest:** Stress and anxiety  **Type of health impacts measured:**  **Physical** –  **Mental** – Trauma  **Social** –  **Economic –** supports the finding that those with lower income are more likely to have developed mental health conditions. | **Main finding:**  Two thirds of respondents did not report any impact on household mental health. Same for depression.  Multiple characteristics needed to predict mental health deterioration. Income and relocation are most strongly predictive factors.  Those with lower income levels are likely to experience severe mental health deterioration after a flood – lowest income 8 times more likely than highest income.  Those who had to relocate for over 6 months were 6 times more likely to experience mental health issues than those not relocated.  **Additional finding:**  60% had anxiety when it rains. 40% reported often experiencing stress.  Characteristics of the flood event are also associated with severity of mental health deterioration, e.g. depth of floodwater. Those flooded at <1m are one third as likely to experience severe mental health deterioration as those with >1m flooding.  Actions taken before the flood to reduce damage is negatively correlated with mental health impact. Implementing mitigation measures reduced the incidence of severe mental health deterioration by four fifths.  The authors suggest this may indicate that a feeling of control over damage could boost mental resilience.  Post flood mitigation measures also positively correlated with mental health issues. Suggestive of experiential learning of flooded households, motivated by avoiding future flooding.  Of post flood stressors, displacement is most strongly correlated with mental health issues.  This is 6 years post event, so it is expected that the proportion affected is much lower than that seen in 1 year follow up studies.  **Recommendations:**  Research the relationship between income levels and reported mental health deterioration. Is it due to resource constraints, or a more general lack or resilience related to low income?  Research to explore the potential of complementing experiential learning with targeted mitigation support to build resilience, especially in communities likely to experience further flooding.  **Sensitivity analysis results (for economic evaluations):** |
| 10. | Mason, V., Andrews, H., & Upton, D. (2010). The psychological impact of exposure to floods. *Psychology, health & medicine*, *15*(1), 61-73. DOI: [**10.1080/13548500903483478**](https://doi.org/10.1080/13548500903483478)  (Mason et al., 2010)  **Country:** UK (Nation countries not specified)  **Aim:** To assess the psychological impact of widespread flooding and to identify risk factors for the development of psychological sequelae in a large population of adults. | **Intervention:** No intervention.  **Dates of data collection:** 5 months after the flood.  **Intervention recipients and sample size:** Overall, 444 (of 3242) completed questionnaires were returned.  **Setting:** UK  **Delivery mode (e.g., remotely online, in person):** Postal questionnaire.  **Intervention deliverers:** No intervention  **Timing and duration:** No intervention  **Intervention description:** No intervention | **Study type:** A cross-sectional study (postal survey).  **Length of follow-up:** No-follow-up.  **Type of economic evaluation/cost analysis:** N/A  **Perspective of analysis:** N/A  **Currency and cost year:** N/A  **Discounting:** N/A  **Sensitivity analysis: N/A**  **Quality appraisal:** High | **Outcome/s of interest:**   - **PTSD** - **Anxiety** - **Depression**   **Types of costs measured:** No costs measured. | **Main finding:** Females had higher mean scores on PTSD, anxiety and depression than males. Most frequently reported coping strategies were rational, detached, and avoidant, with the least frequent being emotional coping. Having to vacate home following flood, previous experience of flooding and poor health was associated with greater psychological distress.  **Additional finding:** Detached coping appeared to be related to less distress. Although it is not possible to determine whether the symptoms were a direct consequence of the flood, symptoms of distress are a significant issue amongst communities affected by environmental events warranting further attention to prevent chronic distress.  **Sensitivity analysis results (for economic evaluations):** No economic analysis conducted. |
| 11. | Medd, W., Deeming, H., Walker, G., Whittle, R., Mort, M., Twigger-Ross, C., Walker, M., Watson, N., & Kashefi, E. (2014). The flood recovery gap: A real-time study of local recovery following the floods of June 2007 in Hull, North East England. Journal of Flood Risk Management, 8(4), 315–328. <https://doi.org/10.1111/jfr3.12098>  (Medd et al., 2014)  **Country:**  England, UK  **Aim:** To bring attention to in-depth research on the processes of recovery and the challenges of addressing the ‘flood recovery’ gap. | **Intervention:** No intervention.  **Dates of data collection:**  2007 – 2008  **Population and sample size:**  44 ‘diarists’, 42 flood affected, 2 frontline workers  **Setting:** Hull, Northeast England  **Delivery mode (e.g., remotely online, in person):** In-person and remote.  **Intervention deliverers:** No intervention.  **Timing and duration:**  18 months.  **Intervention description:** No intervention. | **Study type:** Qualitative. Longitudinal case study. Grounded theory approach to data analysis. Diaries completed by participants over 18-month period. Semi structured interviews at the start. Quarterly group discussions.  **Length of follow-up:** 18 months post flood.  **Type of economic evaluation/cost analysis:** No economic analysis.  **Perspective of analysis:** N/A  **Currency and cost year:** N/A  **Discounting:** N/A  **Sensitivity analysis:** N/A  **Quality appraisal:** Moderate | **Outcome/s of interest:**  Social health  **Types of costs measured:** No costs measured.  **Type of health impacts measured:**  **Physical** – flood recovery process makes day to day illnesses harder to deal with, and conversely being ill exaggerates the stress of the recovery process.  **Mental** – Ongoing stress, which does not necessarily follow a straight-line trajectory in a positive direction.  **Social** – Relationships with family – recognise importance of family support, but also the difficulties faced in family life can be harder to deal with when people have lower reserves.  **Economic** Financial difficulties exacerbated by flooding, or new onset financial difficulties. | **Main finding:**  The ‘recovery gap’ refers to the longer process of recovery, between the points where public authority support diminishes and a less well-defined private sector agencies start. It is often not the floods themselves, but the recovery process that people find difficult to deal with: project managing, 'fighting’, loss of treasured possessions, ‘strip-out’ of the home.  **Additional finding:**  Recovery punctuated by highs and lows, related to personal life events, and experience with organisations. Intangible aspects of flood damage are impossible to quantify. Current lack of space for victims to talk and share experiences, be heard, and listen to key representatives from public and private sector organisations.  **Sensitivity analysis results (for economic evaluations):** No economic analysis conducted. |
| 12. | Mehring, P., Geoghegan, H., Cloke, H. L., & Clark, J. M. (2023). The F word: The experiential construction of flooding in England. *Emotion, Space and Society*, *48*, 100966. DOI: [**10.1016/j.emospa.2023.100966**](https://doi.org/10.1016/j.emospa.2023.100966)  (Mehring et al., 2023)  **Country:** England, UK  **Aim:** To challenge the basic definitions of flooding and the approach that English policy takes in managing it. | **Intervention:** No intervention.  **Dates of data collection:** December 2018 to June 2019.  **Intervention recipients and sample size:** 20 individuals took part in the research.  **Setting:** Face-to-face, via the phone or online.  **Delivery mode (e.g., remotely online, in person):** No intervention.  **Intervention deliverers:** No intervention.  **Timing and duration:** No intervention.  **Intervention description:** No intervention. | **Study type:** Qualitative.  **Length of follow-up:** No follow-up.  **Type of economic evaluation/cost analysis:** No economic analysis.  **Perspective of analysis:** N/A  **Currency and cost year:** N/A  **Discounting:** N/A  **Sensitivity analysis:** No  **Quality appraisal:** High | **Outcome/s of interest:**   - **Emotional impacts** - **Anxiety**   **Types of costs measured:** No costs measured. | **Main finding:** A flood brings with it a tidal wave of emotion. The stress and anguish of water flooding your home, sadness at the loss of precious personal belongings, the fear of rain, anxiety about the return of the flood and the ongoing emotional labour that is required in learning to live at risk of flooding.  **Additional finding:** This research clearly shows that retaining current flood risk management policy means that many thousands of households will suffer the long-term emotional impacts of flooding, unsupported and un-helped, with no-one to provide the hope they need to regain a good quality of life.  **Sensitivity analysis results (for economic evaluations):** No economic analysis conducted. |
| 13. | Milojevic, A., Armstrong, B., & Wilkinson, P. (2017). Mental health impacts of flooding: a controlled interrupted time series analysis of prescribing data in England. *J Epidemiol Community Health*. DOI: [**10.1136/jech-2017-208899**](https://doi.org/10.1136/jech-2017-208899)  (Milojevic et al., 2017)  **Country:** England, UK  **Aim:** To investigate prescription records for drugs used in the management of common mental disorder among primary care practices located in the vicinity of recent large flood events**.** | **Intervention:** Medications for mental disorders (prescriptions).  **Dates of data collection:** England, 2011–2014.  **Intervention recipients and sample size:** N=930 GP practices by flood areas in England.  **Setting:** England.  **Delivery mode (e.g., remotely online, in person):** Secondary data.  **Intervention deliverers:** No intervention.  **Timing and duration:** No intervention.  **Intervention description:** No intervention. | **Study type:** Secondary data (prescription data).  **Length of follow-up:** No follow-up.  **Type of economic evaluation/cost analysis:** No economic analysis.  **Perspective of analysis:** N/A  **Currency and cost year:** N/A  **Discounting:** N/A  **Sensitivity analysis:** No  **Quality appraisal:** Moderate | **Outcome/s of interest:** Antidepressant drugs on prescription.  **Types of costs measured:** No costs measured. | **Main finding:** This study suggests an increase in prescribed antidepressant drugs in the year after flooding in primary care practices close to recent major floods in England.  **Sensitivity analysis results (for economic evaluations):** No economic analysis conducted. |
| 14. | Milojevic, A., Armstrong, B., Kovats, S., Butler, B., Hayes, E., Leonardi, G., Murray, V., & Wilkinson, P. (2011). Long-term effects of flooding on mortality in England and Wales, 1994-2005: Controlled interrupted time-series analysis. Environmental Health: A Global Access Science Source, 10(1), 1–9. https://doi.org/10.1186/1476-069X-10-11  (Milojevic et al., 2011)  **Country:** England and Wales, UK  **Aim:** To explore the methods for quantifying long-term health effects of flooding by analysis of routine mortality registrations in England and Wales. | **Intervention:** No intervention.  **Dates of data collection:** 1994-2005.  **Intervention recipients and sample size:** 319 recorded floods.  **Setting:** England and Wales.  **Delivery mode (e.g., remotely online, in person):** N/A  **Intervention deliverers:** N/A  **Timing and duration:** N/A  **Intervention description:** N/A | **Study type:**  **Length of follow-up:** No follow-up.  **Type of economic evaluation/cost analysis:** No economic analysis.  **Perspective of analysis:** N/A  **Currency and cost year:** N/A  **Discounting:** N/A  **Sensitivity analysis:** No.  **Quality appraisal:** Moderate | **Outcome/s of interest:**  Mortality after floods.  **Types of costs measured:** No costs measured. | **Main finding:** Among the 319 recorded floods, there were 771 deaths in the year before flooding and 693 deaths in the year after (post-/pre-flood ratio of 0.90, 95% CI 0.82, 1.00). This ratio did not vary substantially by age, sex, population density or deprivation. A similar post-flood ‘deficit’ of deaths was suggested by the analyses based on observed/expected deaths  **Sensitivity analysis results (for economic evaluations):** No economic analysis conducted. |
| 15. | Mulchandani, R., Armstrong, B., Beck, C. R., Waite, T. D., Amlôt, R., Kovats, S., ... & Oliver, I. (2020). The English National Cohort Study of Flooding & Health: psychological morbidity at three years of follow up. *BMC public health*, *20*(1), 1-7. Doi: [**10.1186/s12889-020-8424-3**](https://doi.org/10.1186%2Fs12889-020-8424-3)  (Mulchandani et al., 2020)  **Country:** England, UK  **Aim:** To investigate 3 years of data from the English National Study of Flooding and Health. | **Intervention:** No intervention.  **Dates of data collection:** Data from 2015,2016, 2017.  **Intervention recipients and sample size:** No intervention.  **Setting:** England, UK.  **Delivery mode (e.g., remotely online, in person):** No intervention.  **Intervention deliverers:** No intervention.  **Timing and duration:** No intervention.  **Intervention description:** No intervention. | **Study type:** Survey.  **Length of follow-up:** This was a follow-up, two years after the original survey started in 2015.  **Type of economic evaluation/cost analysis:** No economic analysis.  **Perspective of analysis:** N/A  **Currency and cost year:** N/A  **Discounting:** N/A  **Sensitivity analysis:** No.  **Quality appraisal:** High | **Outcome/s of interest:** This was a follow-up, two years after the original survey started in 2015.  Flooding can have severe long-lasting consequences on mental health in affected populations**.** Eight hundred nineteen individuals were included in the final analysis – 119 were classified as unaffected, 421 disrupted and 279 flooded. Overall, 5.7% had probable depression, 8.1% had probable anxiety and 11.8% had probable PTSD, with higher prevalence in the flooded group compared with the unaffected group. After adjustment for potential confounders, probable mental health outcomes were higher in the flooded group compared to the unaffected group, significantly for probable depression (aOR 8.48, 95% CI 1.04–68.97) and PTSD (aOR 7.74, 95% CI 2.24–26.79)  **Types of costs measured:** No costs measured. | **Main finding:** Eight hundred nineteen individuals were included in the final analysis – 119 were classified as unaffected, 421 disrupted and 279 flooded. Overall, 5.7% had probable depression, 8.1% had probable anxiety and 11.8% had probable PTSD, with higher prevalence in the flooded group compared with the unaffected group.  **Additional finding:** Of the 569 participants who responded at all 3 years, a significant reduction in prevalence for all probable mental health outcomes was observed in the flooded group.  **Sensitivity analysis results (for economic evaluations):** No economic analysis conducted. |
| 16. | Munro, A., Kovats, R. S., Rubin, G. J., Waite, T. D., Bone, A., Armstrong, B., Beck, C. R., Amlôt, R., Leonardi, G., & Oliver, I. (2017). Effect of evacuation and displacement on the association between flooding and mental health outcomes: a cross-sectional analysis of UK survey data. The Lancet Planetary Health, 1(4), e134–e141. <https://doi.org/10.1016/S2542-5196(17)30047-5>  (Munro et al., 2017)  **Country:** England, UK  **Aim:** To compare the prevalence of depression, anxiety and PTSD between participants displaced by flooding and those flooded but not displaced. | **Intervention:** No intervention.  **Dates of data collection:**  2015 (1 year after flooding).  **Population and sample size:** 622 flooded respondents from a larger sample of all households – Public Health England National Study of Flooding and Health (n =2126). Same data set as used by Waite et al.. (2017)  **Setting: F**lood-affected neighbourhoods in Gloucestershire, Wiltshire, Surrey, Somerset, and Kent  **Delivery mode (e.g., remotely online, in person):** -Recruitment packs sent to 8761 addresses. Questionnaires returned by post or online.  **Intervention deliverers:**  **Timing and duration:**  Single time point  **Intervention description:** No intervention | **Study type:** Quantitative survey.  **Length of follow-up:** No follow-up.  **Type of economic evaluation/cost analysis:** No economic analysis.  **Perspective of analysis:** N/A  **Currency and cost year:** N/A  **Discounting:** N/A  **Sensitivity analysis:** N/A  **Quality appraisal:** High | **Outcome/s of interest:**  Depression, anxiety, and PTSD.  **Types of costs measured:** No costs measured.  **Type of health impacts measured:**  **Physical** –  **Mental** – Depression, anxiety, and PTSD  **Social** –  **Economic** | **Main finding:**  Findings People who were displaced from their homes were significantly more likely to have higher scores on each scale; odds ratio (OR) for depression 1·95 (95% CI 1·30–2·93), for anxiety 1·66 (1·12–2·46), and for post-traumatic stress disorder 1·70 (1·17–2·48) than people who were not displaced. The increased risk of depression was significant even after adjustment for severity of flooding. Scores for depression and post-traumatic stress disorder were higher in people who were displaced and reported receiving no warning than those who had received a warning more than 12 h in advance of flooding (p=0·04 for depression, p=0·01 for post-traumatic stress disorder), although the difference in anxiety scores was not significant.  **Additional finding:**  The amount of warning received showed evidence of being protective against the mental illnesses. Severity of flooding may account for some differences between groups.  **Sensitivity analysis results (for economic evaluations):** No economic analysis conducted. |
| 17. | Paranjothy, S., Gallacher, J., Amlôt, R., Rubin, G. J., Page, L., Baxter, T., Wight, J., Kirrage, D., McNaught, R., & Palmer, S. R. (2011). Psychological impact of Summer 2007 floods in England. BMC Public Health, 11, 145–153. DOI: 10.1186/1471-2458-11-145  (Paranjothy et al., 2011)  **Country:** England, UK  **Aim:** To compare prevalance of 4 mental health symptoms (psychological distress, generalised anxiety disorder, depression and post traumatic stress disorder) between flooded and non-flooded population, and the role of risk factors. | **Intervention:** No intervention.  **Dates of data collection:** 2007-2008  **Population and sample size:**  2265 questionnaires completed.  **Setting:** South Yorkshire and Worcestershire  **Delivery mode (e.g., remotely online, in person):** post, online, telephone or face to face.  **Intervention deliverers:**  Study carried out by Health Protection Agency and local public health departments.  **Timing and duration:** 3-6 months after flooding  **Intervention description:**  Multivariable logistic regression used to describe association between water level in home and mental health symptoms. | **Study type:** Quantitative. Surveys in 2 regions,  **Length of follow-up:** No follow-up.  **Type of economic evaluation/cost analysis:** No economic analysis.  **Perspective of analysis:** N/A  **Currency and cost year:** N/A  **Discounting:** N/A  **Sensitivity analysis:** N/A  **Quality appraisal:** High | **Outcome/s of interest:**  Psychosocial health impact  **Types of costs measured:** None. | **Main finding:**  Mental health symptoms significantly higher in individuals who reported flood water in the home than those who did not, with prevalence increasing with the level of flood water in the home.  **Additional finding:**  Mental health conditions were 3-5 times higher in the more socio-economically deprived Yorkshire study area than in Worcestershire.  Symptoms significantly raised in women, unemployed and those with pre-existing health conditions.  Perceived risk impacts on mental health -beliefs about health effects and financial implications are directly linked to mental health symptoms, with a 2 to 40-fold increase in prevalence.  Loss of essential services also a risk factor, worsening mental health 2-3 fold.  **Sensitivity analysis results (for economic evaluations):** No economic analysis conducted. |
| 18. | Reacher M, McKenzie K, Lane C, Nichols T, Kedge I, Iversen A, Hepple P, Walter T, Laxton C, Simpson J. (2004). Health impacts of flooding in Lewes: a comparison of reported gastrointestinal and other illness and mental health in flooded and non-flooded households. Commun Dis Public Health ;7(1):39-46. PMID: 15137280.  <https://pubmed.ncbi.nlm.nih.gov/15137280/>  **Country:** England, UK  **Aim:**  To investigate independent association between flooding, psychological and physical health. | **Intervention:** N/A  **Dates of data collection:** July-August 2001  **Intervention recipients and sample size:**  227 residents were flooded.  **Setting:** Lewes, Southern England  **Delivery mode (e.g., remotely online, in person):** N/A  **Intervention deliverers:** N/A  **Timing and duration:** N/A  **Intervention description:** N/A | **Study type:**  Cohort study  Interviews and questionnaires for 227 flood exposed individuals and non-flooded households 240 individuals  **Length of follow-up:** N/A  **Type of economic evaluation/cost analysis:** N/A  **Perspective of analysis:** N/A  **Currency and cost year:** N/A  **Discounting:** N/A  **Sensitivity analysis:** N/A  **Quality appraisal:** Moderate | **Outcome/s of interest:**  Psychological and physical health following flooding.  **Types of costs measured:** None. | **Main finding:**  Flooding remained highly significantly associated with psychological distress after adjustment for physical illnesses. Psychological distress may explain some of the excess physical illness reported by flooded adults and possibly by children as well.  **Additional finding:**  flooded was associated with earache p = 0.02  increase in risk of gastroenteritis p = 0.09  **Sensitivity analysis results (for economic evaluations):** N/A |
| 19. | Robin C, Beck C, Armstrong B, Waite TD, Rubin GJ; English National Study of Flooding and Health Study Group; Oliver I. Impact of flooding on health-related quality of life in England: results from the National Study of Flooding and Health. Eur J Public Health. 2020 Oct 1;30(5):942-948. doi: 10.1093/eurpub/ckaa049. <https://pubmed.ncbi.nlm.nih.gov/32227174/>  **Country:** England, UK  **Aim:** The aim of this study was to describe the HRQoL of individuals exposed to flooding following the 2013/14 winter storms in England, using data from the National Study  of Flooding and Health (NSFH), 2- and 3-years post-flooding | **Intervention:** N/A  **Dates of data collection:**  **Intervention recipients and sample size:**  **Setting:** England  **Delivery mode (e.g., remotely online, in person):**  **Intervention deliverers:** N/A  **Timing and duration:** N/A  **Intervention description:** N/A | **Study type:**  Cohort study  Interviews and questionnaires for 227 flood exposed individuals and non-flooded households 240 individuals  **Length of follow-up:** N/A  **Type of economic evaluation/cost analysis:** N/A  **Perspective of analysis:** N/A  **Currency and cost year:** N/A  **Discounting:** N/A  **Sensitivity analysis:** N/A  **Quality appraisal: High** | **Outcome/s of interest:**  Health-related quality of life  **Types of costs measured:** N/A | **Main finding:**  For both 2- and 3-years post-flooding, the median HRQoL scores were lower in the flooded and disrupted  groups, compared with unaffected respondents. A higher proportion of flooded and disrupted respondents  reported HRQoL problems in most dimensions of the EQ-5D-5L, compared with unaffected respondents. In  year 2, independent associations between exposure to flooding and experiencing anxiety/depression [adjusted  odds ratio (aOR) 7.7; 95% CI 4.6–13.5], problems with usual activities (aOR 5.3; 95% CI 2.5–11.9) and pain/discomfort  (aOR 2.4; 95% CI 1.5–3.9) were identified. These problems persisted 3 years post-flooding; associations  between exposure to flooding and experiencing anxiety/depression (aOR 4.3; 95% CI 2.5–7.7), problems with  usual activities (aOR 2.9; 95% CI 1.5–6.1) and pain/discomfort (aOR 2.5; 95% CI 1.5–4.2) were identified.  **Additional finding:**  **Sensitivity analysis results (for economic evaluations):** |
| 20. | Tapsell, S. M., Penning-Rowsell, E. C., Tunstall, S. M., & Wilson, T. L. (2002). Vulnerability to flooding: health and social dimensions. Philosophical Transactions of the Royal Society of London, 1511–1525.DOI: 10.1098/rsta.2002.1013  (Tapsell et al., 2002)  **Country:**  England, UK  **Aim:** To better understand the social effects of flooding – those caused by disruption that do not carry a monetary price tag. | **Intervention:** No intervention, exploratory study. Developed a model of flood impacts, based on 3 social characteristics and 4 financial deprivation indicators - Social Flood vulnerability index (SFVI)  **Dates of data collection:**  2000  **Population and sample size:**  6 focus groups.  **Setting:** Three flood affected villages in England  **Delivery mode (e.g., remotely online, in person):** In-person  **Intervention deliverers:** No intervention.  **Timing and duration:**  Focus groups carried out 3-4 months post flooding.  **Intervention description:** No intervention. | **Study type:** Qualitative. Focus group discussions initially, followed by a quantitative model to predict social vulnerability.  **Length of follow-up:** None  **Type of economic evaluation/cost analysis:** None. However, this paper refers to cost/benefit analysis of flood defences.  **Perspective of analysis:** Those directly affected by flooding.  **Currency and cost year:** N/A  **Discounting:** N/A  **Sensitivity analysis:** N/A  **Quality appraisal:** Moderate | **Outcome/s of interest:**  Social health  **Types of costs measured:**  **Type of health impacts measured:**  **Physical** – self reported physical impacts.  **Mental** – Trauma effects, ranging in severity. Not all mental health impacts are severe enough to have a diagnosis, yet the effects are still impacting on lives, e.g. sense of loss, helplessness, worry about perceived risks.  **Social** – ‘Flood victim’, ‘feel like a leper’, change in neighbourhood, meaning of home, media intrusion, ‘lost time’ – feeling that the flooding and recovery has taken over their lives.  **Economic** Financial deprivation recognised as predisposing factor in vulnerability to flooding. | **Main finding:**  Social factors thought to predispose people to worse effects, and therefore used in the SFVI are elderly (75+), lone parents, pre-existing health problems, and financial deprivation.  **Additional finding:**  Assessments of the effect of flood defence measures on reducing impacts are flawed if only monetary losses are used in cost/benefit analysis for investment appraisals.  Psychosocial consequences from the trauma of flooding go beyond diagnosable mental health conditions.  Women more affected by flooding (physical and mental) than men.  **Sensitivity analysis results (for economic evaluations):** No economic analysis conducted. |
| 21. | Waite, T. D., Chaintarli, K., Beck, C. R., Bone, A., Amlôt, R., Kovats, S., Reacher, M., Armstrong, B., Leonardi, G., Rubin, G. J., & Oliver, I. (2017). The English national cohort study of flooding and health: cross-sectional analysis of mental health outcomes at year one. BMC Public Health, 17(1), 1–9. https://doi.org/10.1186/s12889-016-4000-2  (Waite et al., 2017)  **Country:**  UK, England  **Aim:** To describe the prevalence of probable depression, anciety and PTSD at one year among individuals exposed to flooding or disruption from flooding compared to those unexposed.  To identify personal and sociodemographic characteristics associated with psychological morbidity. | **Intervention:** No intervention.  **Dates of data collection:**  January to March 2015, one year post flooding.  **Population and sample size:** 2006 participants analysed. 23% response to invitation.  **Setting:** Wiltshire, Surrey, Gloucestershire, Sedgemoor, South Somerset, and Tonbridge and Malling.  Included some of the most affluent areas in the UK  **Delivery mode (e.g., remotely online, in person):** online or by post  **Intervention deliverers:**  **Timing and duration:**  Cross section  **Intervention description:** Use of validated tools in survey. | **Study type:** Quantitative. Cross sectional analysis nested within a wider cohort study - Public Health England National Study of Flooding and Health (n =2126). Same data set as used by Munro et al.. (2017)  **Length of follow-up:** None  **Type of economic evaluation/cost analysis:** None  **Perspective of analysis:** Post code areas affected by flooding  **Currency and cost year:** N/A  **Discounting:** N/A  **Sensitivity analysis:** N/A  **Quality appraisal:** High | **Outcome/s of interest:** Anxiety, depression, and PTSD.  **Types of costs measured:** No costs were measured.  **Type of health impacts measured:**  **Physical** –  **Mental** – Severity of flooding associated with increased odds of psychological morbidity.  **Social** – Neighbourhood effect – increased psychological morbidity seen even in those whose homes were unaffected by the floods. Much of the communities were still affected by disruption to utilities, transport, education, work and access to health and social services.  **Economic** Financial. | **Main finding:** 2126 people (23%) responded. The prevalence of psychological morbidity was elevated amongst flooded participants ([n = 622] depression 20.1%, anxiety 28.3%, PTSD 36.2%) and disrupted participants ([n = 1099] depression 9.6%, anxiety 10.7% PTSD 15.2%). Flooding was associated with higher odds of all outcomes (adjusted odds ratios (aORs), 95% CIs for depression 5.91 (3.91–10.99), anxiety 6.50 (3.77–11.24), PTSD 7.19 (4.33–11.93)). Flooded participants who reported domestic utilities disruption had higher odds of all outcomes than other flooded participants, (aORs, depression 6.19 (3.30–11.59), anxiety 6.64 (3.84–11.48), PTSD 7.27 (4.39–12.03) aORs without such disruption, depression, 3.14 (1.17–8.39), anxiety 3.45 (1.45–8.22), PTSD 2.90 (1.25–6.73)). Increased floodwater depth was significantly associated with higher odds of each outcome. Disruption without flooding was associated with borderline higher odds of anxiety (aOR 1.61 (0.94–2.77)) and higher odds of PTSD 2.06 (1.27–3.35)) compared with unaffected participants. Disruption to health/social care and work/education was also associated with higher odds of psychological morbidity.  **Additional finding:**  Effects of flooding extend beyond those immediately affected. At a community level, reinstating access to transport, education, work and health and social care services as soon as possible may be protective against mental health morbidity.  **Sensitivity analysis results (for economic evaluations):** No economic analysis conducted. |

**END**
