## Supplementary File 3 for "Extreme weather events in the UK and resulting public health outcomes"

### Supplementary file 3 - Evidence table for Extreme Weather Events (EXE) Systematic review (SR) included papers: Extreme heat

| **Citation (Country)**  **Aim** | **Intervention details** | **Study characteristics, health economics methods and quality appraisal** | **Outcome and costs measured** | **Main Findings** |
| --- | --- | --- | --- | --- |
| Alahmad, B., Khraishah, H., Royé, D., Vicedo-Cabrera, A. M., Guo, Y., Papatheodorou, S. I., Achilleos, S., Acquaotta, F., Armstrong, B., Bell, M. L., Pan, S. C., De Sousa Zanotti Stagliorio Coelho, M., Colistro, V., Dang, T. N., Van Dung, D., De’ Donato, F. K., Entezari, A., Guo, Y. L. L., Hashizume, M., … Koutrakis, P. (2023). Associations Between Extreme Temperatures and Cardiovascular Cause-Specific Mortality: Results From 27 Countries. Circulation, 147(1), 35–46. <https://doi.org/10.1161/CIRCULATIONAHA.122.061832>  (Alahmad et al., 2023)  **Country:**  Worldwide including the UK  **Aim:** To assemble a database of daily counts of specific cardiovascular causes of death from 567 cities in 27 countries across 5 continents in overlapping periods ranging from 1979 to 2019. | **Intervention:** No intervention.  **Dates of data collection:** 1979 to 2019.  **Population and sample size:**  The analyses included deaths from any cardiovascular cause (32 154 935), ischemic heart disease (11 745 880), stroke (9 351 312), heart failure (3 673 723), and arrhythmia (670 859).  **Setting:** Worldwide  **Delivery mode (e.g., remotely online, in person):** N/A  **Intervention deliverers:** N/A  **Timing and duration:** N/A  **Intervention description:** N/A | **Study type:** Quantitative.  **Length of follow-up:** No follow-up.  **Type of economic evaluation/cost analysis:** No economic analysis.  **Perspective of analysis:** N/A  **Currency and cost year:** N/A  **Discounting:** N/A  **Sensitivity analysis:** N/A | **Outcome/s of interest:**  Mortality rates in extreme hot and cold temperatures.  **Types of costs measured:** No costs measured. | **Main finding:**  At extreme temperature percentiles, heat (99th percentile) and cold (1st percentile) were associated with higher risk of dying from any cardiovascular cause, ischemic heart disease, stroke, and heart failure as compared to the minimum mortality temperature, which is the temperature associated with least mortality.  **Additional finding:**  Across a range of extreme temperatures, hot days (above 97.5th percentile) and cold days (below 2.5th percentile) accounted for 2.2 (95% empirical CI [eCI], 2.1–2.3) and 9.1 (95% eCI, 8.9–9.2) excess deaths for every 1000 cardiovascular deaths, respectively. Heart failure was associated with the highest excess deaths proportion from extreme hot and cold days with 2.6 (95% eCI, 2.4–2.8) and 12.8 (95% eCI, 12.2–13.1) for every 1000 heart failure deaths, respectively.  **Sensitivity analysis results (for economic evaluations):**  **Recommendations:** No economic analysis conducted. |
| Arbuthnott KG, Hajat S. The health effects of hotter summers and heat waves in the population of the United Kingdom: a review of the evidence. Environ Health. 2017 Dec 5;16(Suppl 1):119. doi: 10.1186/s12940-017-0322-5.  (Arbuthnott & Hajat, 2017)    **Country:** UK  **Aim:** to bring together evidence from epidemiological studies and health impact assessments to provide an overview of what is known about current and projected effects of heat on population level health in the UK. | **Intervention:**  Not an intervention study  **Dates of data collection:**  N/A  **Intervention recipients and sample size:** N/A  **Setting:** N/A  **Delivery mode (e.g., remotely online, in person):** N/A  **Intervention deliverers:** N/A  **Timing and duration:** N/A  **Intervention description:** N/A | **Study type:**  Literature Review  **Length of follow-up:** N/A  **Type of economic evaluation/cost analysis:** N/A  **Perspective of analysis:** N/A  **Currency and cost year:** N/A  **Discounting:** N/A  **Sensitivity analysis:** N/A | **Outcome/s of interest:**  Effect of heat on health   - mortality - mortality displacement - morbidity - other indicators of health and wellbeing - work productivity   Distribution of health impacts by   - geographical/regional differences - age - sex   **Types of costs measured:**  N/A | **Main finding:** 1) UK studies illustrating an increase in heat-related mortality occurring at temperatures above threshold values, with respiratory deaths being more sensitive to heat than deaths from cardiovascular disease (although the burden from cardiovascular deaths is greater in absolute terms).  2)The relationship between heat and other health outcomes are less consistent.  3)Within the UK, the main populations who are vulnerable to heat were the older populations; those with certain co-morbidities and those living in Greater London, the Southeast and Eastern regions.  **Additional finding:**  key gaps in knowledge: how urbanization and population  adaptation to heat will affect health impacts, and current and future strategies for effective, sustainable  and equitable adaptation to heat.  **Sensitivity analysis results (for economic evaluations):**  N/A |
| Berger, S. E., Ordway, M. R., Schoneveld, E., Lucchini, M., Thakur, S., Anders, T., Natale, L., & Barnett, N. (2023). The impact of extreme summer temperatures in the United Kingdom on infant sleep: Implications for learning and development. Scientific Reports, 13(1), 1–8. https://doi.org/10.1038/s41598-023-37111-2  (Berger et al., 2023)  **Country:** England, UK  **Aim:** To assess the impact of unusually high nighttime temperatures on infant sleep. | **Intervention:** No intervention.  Users of the Nanit baby monitor received an email invitation to participate in a study investigating the association between sleep and climate change. Users who agreed to participate in the study gave their informed consent to share objective sleep data collected in the summer of 2022 using a computer-vision algorithm for research purposes.  **Dates of data collection:**  Summer 2022, during which three heatwaves occurred.  **Population and sample size:**  Parents of 413 infants (51% girls, 37% boys, 12% not reported; Mean age as of June 1, 2022=12.22 months, Median age=11 months, SD=5.48 months, Range=2 months to 33 month)  **Setting:** Greater London, England  **Delivery mode (e.g., remotely online, in person):** -No intervention.  **Intervention deliverers:** No intervention.  **Timing and duration:** N/A  **Intervention description:** No intervention. | **Study type:** Quantitative. exploratory.  Sleep–wake pattern.  Changepoint analysis used to identify abrupt mean changes in time series data.  **Length of follow-up:** No follow-up.  **Type of economic evaluation/cost analysis:** No economic analysis.  **Perspective of analysis:** N/A  **Currency and cost year:** N/A  **Discounting:** N/A  **Sensitivity analysis:** N/A | **Outcome/s of interest:**  (1) total sleep time (TST; total minutes scored as sleep within the night sleep period); (2) minutes to sleep onset (MSO; latency until the first minute of 5 consecutive minutes of sleep after infants are placed in the crib); (3) sleep efficiency (SE; the proportion of time the infant spent asleep over the time the infant spent in the crib during the predefined nighttime sleep period) (4) number of night wakings (NW); and (5) number of parental visits to the crib (Visits).  **Types of costs measured:** No costs measured.  **Limitations**  Competing interests – two of the authors are employed by Nanit, the brand of baby monitor used in the study.  Used max daytime temperatures and not ambient room temperature.  Daytime naps were not captured.  Self-selecting sample – only parents who could afford to buy the Nanit baby monitor. | **Main finding:** Deviations found in all sleep metrics. Infants slept less, took longer to fall asleep, had less efficient sleep, more night awakenings, and more parent visits to the crib during heat wave periods. Statistical data not presented in the paper – supplementary material available on request.  **Additional finding:**  The timing of sleep disruptions was associated with the absolute value of the temperature, not the relative value that defines a heatwave. Sleep was not impacted when temps stayed below 88 degrees Fahrenheit (31.1 Celsius) but was significantly negatively affected when temperatures reached over 100 degrees (37.8 Celsius).  Infants slept on average 20 minutes less during the 2^nd^ heatwave. Previous research suggests even small amounts of sleep deprivation cause detrimental effects.  **Sensitivity analysis results (for economic evaluations):**  **Recommendations:** Further research should look at prospective behavioural studies of problem solving or memory consolidation before, during, and after extreme temperature events. |
| Bryan, K., Ward, S., Roberts, L. et al.. The health and well-being effects of drought: assessing multi-stakeholder perspectives through narratives from the UK. Climatic Change 163, 2073–2095 (2020). <https://doi.org/10.1007/s10584-020-02916-x>  (Bryan et al., 2020)  **Country:** UK (regions in Scotland, England and Wales)  **Aim:** to assess people’s narratives of drought on health and well-being in the UK using source-receptor-impact framing | **Intervention:** Not an intervention study  **Dates of data collection:**  N/A  **Intervention recipients and sample size:**  N= 41  **Setting:**  semi-structured and narrative interviews, story sharing and  creation of short video stories and collection of short audio-recorded stories (micro-narratives) at  events such as festivals and river walks to build a repository of stories  **Delivery mode (e.g., remotely online, in person):** In person  **Intervention deliverers:** N/A  **Timing and duration:** N/A  **Intervention description:** N/A  **Quality appraisal:** Moderate | **Study type:**  Qualitative and narrative  **Length of follow-up:**  N/A  **Type of economic evaluation/cost analysis**  N/A  **Perspective of analysis:**  Perspective of drought-related narratives of participants from diverse backgrounds  **Currency and cost year:**  N/A  **Discounting:**  N/A  **Sensitivity analysis:**  N/A | **Outcome/s of interest:**  Identified sources of drought-health effects   - Water quantity, hygiene and sanitation - Water quality (PWS users and at-risk groups; ecosystems and human health) - Food security - Air quality - Heatwave - Vectors   In addition, a major outcome of this research was the consensus on the perceived exposure of at-risk groups as likely early receptors of the health and well-being impacts of drought, and the need for targeted action.  **Types of costs measured:**  N/A | **Main finding:**  1) Drought can present perceived health and well-being effects through reduced water quantity, water quality, compromised hygiene and sanitation, food security, and air quality  2) Heatwave associated with drought was also identified as a source of health effects through heat and wildfire, and drought-related vectors.  3) Drought was viewed as potentially attributing both negative and positive effects for physical and mental health, with emphasis on mental health.  **Additional finding:**  Two recurring themes in the UK narratives were the health consequences of drought for ‘at-risk’ groups and the need to target them, and that drought in a changing climate presented potential health implications for at-risk groups.  **Sensitivity analysis results (for economic evaluations):**  N/A |
| Cruz, J., White, P. C. L., Bell, A., & Coventry, P. A. (2020). Effect of extreme weather events on mental health: A narrative synthesis and meta-analysis for the UK. International Journal of Environmental Research and Public Health, 17(22), 1–17. <https://doi.org/10.3390/ijerph17228581>  (Cruz et al., 2020)  **Country:** UK  **Aim:** To quantify the prevalence and describe the causes of common mental health problems in populations exposed to extreme weather events in the UK | **Intervention:** No intervention.  **Dates of data collection:**  December 2019  Search from inception to 12 December 2019.  **Population and sample size:**  17 studies were included. Only 1 paper was about heatwaves, the others were flooding. Four of the included studies included in meta-analyses to determine the point prevalence of common mental health problems in the period within 12 months following extreme weather events.  **Setting:** UK (nationwide)  **Delivery mode (e.g., remotely online, in person):** -No intervention.  **Intervention deliverers:** No intervention.  **Timing and duration:**  Search carried out from inception to 12 December 2019.  **Intervention description:** No intervention. | **Study type:**  Systematic review to quantify the prevalence and describe the causes of common mental health problems in populations exposed to extreme weather events in the UK.  Narrative analysis.  Meta analysis of 4 studies (all included in our review): (French et al.. 2019), Graham et al.. (2019), Mason et al. (2010) and Munro et al.. (2017).  **Length of follow-up:** No follow-up.  **Type of economic evaluation/cost analysis:** No economic analysis.  **Perspective of analysis:** N/A  **Currency and cost year:** N/A  **Discounting:** N/A  **Sensitivity analysis:** N/A | **Outcome/s of interest:**  Anxiety  Depression  PTSD  **Types of costs measured:** No costs measured.  **Limitations:**  Studies selected and data extracted by only one reviewer.  Only assessed Flooding and mental health, as only one heat paper identified.  Further research recommendations  insufficient evidence relating to other types of incidents, including heatwaves. Additionally, we found that people from more deprived backgrounds who rented property had poorer psychological health after flooding. Future qualitative research could address questions about the differential impact of flooding among populations in relation to income, home ownership and ethnicity. | **Main finding:**  Meta analysis:  The point prevalence was 19.8% for anxiety (k = 4; n = 1458; 95% CI 7.42 to 32.15), 21.35% for depression (k = 4; n = 1458; 95% CI 9.04 to 33.65) and 30.36% for PTSD (k = 4; n = 1359; 95% CI 11.68 to 49.05).  Narrative analysis identified 6 themes:  mental health morbidity; physical health and longer-term effects on mental health; characteristics of the flood (e.g., increase in water depth); flood warning; displacement and loss of sense of place; and socio-economic impact  **Additional finding:**  Displacement from home was a key factor in the narrative around the impact of flooding on mental health.  At the community level, some studies reported a negative impact, with disrupted activities and loss of community spirit, while others reported increased community resilience and reduced psychological distress owing to social cohesion and collective efficacy to combat the effects of the floods.  Sociodemographic evidence – women more affected, ethnic minorities more affected, age <65 more affected.  Those with lower income levels more affected.  The loss of a sense of place that stemmed from displacement relates to the concept of place attachment, which is a concept that describes the psychological and emotional bonds between people and places. Disruption to these bonds can lead to solastalgia, which refers to distress caused by environmental degradation and loss of home and belongings.  **Sensitivity analysis results (for economic evaluations):** No economic analysis conducted.  **Recommendations:**  Psychological therapies should be made available at a larger scale within flooded communities, not only to those meeting clinical diagnosis thresholds.  Preventative approaches to build resilience  Increase mental health literacy, so people know when they need to seek help.  calls for novel approaches that draw on systems thinking that can propose integrated solutions to address the connections between climate change and climate mitigation and the persistent social determinants of mental ill health |
| Curtis, S., Fair, A., Wistow, J., Val, D. V., & Oven, K. (2017). Impact of extreme weather events and climate change for health and social care systems. Environmental Health: A Global Access Science Source, 16(Suppl 1). <https://doi.org/10.1186/s12940-017-0324-3>  (Curtis et al., 2017)  **Country:** England, UK  **Aim:** To assess the main policy relevant messages from research relevant to health and care systems the UK.  Two sets of questions:  What is the evidence concerning observed impact of these events on service operability and access and on pressure of service demand on the care system; second, what is the evidence regarding potential future impacts of extreme weather on health services in light of projected climate change (as projected up to 2050–80) and potential for adaptation of health and social care infrastructure in the context of climate change.  What is the evidence concerning observed impact of these events on service operability and access and on pressure of service demand on the care system;  what is the evidence regarding potential future impacts of extreme weather on health services considering projected climate change (as projected up to 2050–80) and potential for adaptation of health and social care infrastructure in the context of climate change | **Intervention:** No intervention.  **Dates of data collection:**  2014-2015.  **Population and sample size:**  A ‘structured’ review approach, adapting the procedures required for formal systematic reviews. Not a formal systematic review as only one reviewer.  **Setting:** Nationwide  **Delivery mode (e.g., remotely online, in person):** -No intervention.  **Intervention deliverers:** No intervention.  **Timing and duration:**  Review period 2010 - 2017  **Intervention description:** No intervention. | **Study type:** Quantitative. Cross sectional postal survey.  **Length of follow-up:** No follow-up.  **Type of economic evaluation/cost analysis:** No economic analysis.  **Perspective of analysis:** N/A  **Currency and cost year:** N/A  **Discounting:** N/A  **Sensitivity analysis:** N/A | **Outcome/s of interest:**  Observed impacts of extreme weather on health service infrastructure.  Potential future impacts of extreme weather on health services in light of projected climate change and potential for adaptation.  Potential for adaptation of infrastructure design and practice for improved preparedness and performance during extreme weather.  **Types of costs measured:** No costs measured. | **Main finding:**  Observed impacts of extreme weather on health service infrastructure – divided into themes of extreme heat (thermal comfort of buildings, increased presentation at GPs and worsening of cardiovascular conditions), extreme cold (transport difficulties and increased presentation of respiratory illness and accidents) and flooding (damage to facilities, power and water supply interruptions, patient records, ambulance access, drowning, injury, toxicity, and longer term mental and physical illnesses.  Potential future impacts of extreme weather on health services in light of projected climate change and potential for adaptation.  Difficulties in making projection models and analysing cost-benefit ratio when the future is so uncertain.  Potential for adaptation of infrastructure design and practice for improved preparedness and performance during extreme weather  Building design to improve thermal comfort – planning future health service buildings, and optimising current buildings.  Wider infrastructure – ‘whole systems’ approach required. Social infrastructure – need to understand individual vulnerabilities to modify collective social and institutional factors. Enhanced risk governance – improvements in evaluation of and response to risks – adaptive capacity index suggested.  **Additional finding:**  Findings draw attention to inequalities between socio-economic groups in exposures to and impacts of EWEs.  Syndromic surveillance becoming popular as a method of tracking impacts of EWEs.  **Sensitivity analysis results (for economic evaluations):** No economic analysis conducted. They refer to two studies which have begun to explore cost effectiveness of measures to make health and social care more resilient to extreme weather –  “Attempts to model the cost effectiveness of implementing strategies such as cold weather plans for the NHS are limited by lack of detailed information on the extent to which these plans are being implemented and targeted towards those most in need” .Public Health England Strategic Health Asset Planning and Evaluation. 2015. 19.7.15; Available from: <https://shape.phe.org.uk/>.  Also, Chalabi Z, et al.. Evaluation of the cold weather plan for England: modelling of cost-effectiveness. Public Health. 2016;137:13–9  **Recommendations:**  Preparedness and emergency response strategies call for action extending beyond the emergency response systems, to include health and social care providers. |
| Finlay SE, Moffat A, Gazzard R, Baker D, Murray V. Health impacts of wildfires. PLoS Curr. 2012 Nov 2;4:e4f959951cce2c. doi: 10.1371/4f959951cce2c  (Finlay et al., 2012)  **Country:** England, UK  **Aim:**  to collate and review the evidence regarding human health impacts from global wildfire experience. | **Intervention:**  Not an intervention study  **Dates of data collection:**  **Intervention recipients and sample size:** N/A  **Setting:**  A literature review of current evidence about the health effects of wildfires from the UK standpoint.  **Delivery mode (e.g., remotely online, in person):**  N/A  **Intervention deliverers:**  N/A  **Timing and duration:**  N/A  **Intervention description:**  N/A | **Study type:**  Literature review  **Length of follow-up:**  N/A  **Type of economic evaluation/cost analysis**  N/A  **Perspective of analysis:**  Applying the Source-Pathway-Receptor Exposure Model to assess health threats of wildfires  **Currency and cost year:**  N/A  **Discounting:**  N/A  **Sensitivity analysis:**  N/A | **Outcome/s of interest:**  Impact of acute and chronic chemical exposures on health   - Toxicology of wildfire smoke - Effects of particulate matter in wildfire smoke - Respiratory symptoms - Burns - Heat induced illness - Cardiovascular effects - Ophthalmic effects - Psychological effects - Paediatric psychiatric morbidity   **Types of costs measured:**  N/A | **Main finding:**  1) human health can be severely affected by wildfires  2) Certain populations are particularly vulnerable.  3) Wood smoke has high levels of particulate matter and toxins. Respiratory morbidity predominates, but cardiovascular, ophthalmic and psychiatric problems can also result.  4) Severe burns resulting from direct contact with the fire require care in special units and carry a risk of multi – organ complications.  5) The wider health implications from spreading air, water and land pollution are of concern.  6) Access to affected areas and communication with populations living within them is crucial in mitigating risk.  **Additional finding:**  N/A  **Sensitivity analysis results (for economic evaluations):**  N/A |
| Green, H. K., Andrews, N., Armstrong, B., Bickler, G., & Pebody, R. (2016). Mortality during the 2013 heatwave in England - How did it compare to previous heatwaves? A retrospective observational study. Environmental Research, 147, 343–349. <https://doi.org/10.1016/j.envres.2016.02.028>  (Green et al., 2016)  **Country:** England, UK  **Aim:** To estimate excess mortality rates over several heatwave periods in England. | **Intervention:** No intervention.  **Dates of data collection:**  1 June to 15 September 2013.  **Population and sample size:** 301 deaths in all areas of England.  **Setting:** England, UK  **Delivery mode (e.g., remotely online, in person):** N/A  **Intervention deliverers:** N/A  **Timing and duration:** N/A  **Intervention description:** N/A | **Study type:** A retrospective observational study.  **Length of follow-up:** No follow-up.  **Type of economic evaluation/cost analysis:** No costs measured.  **Perspective of analysis:** N/A  **Currency and cost year:** N/A  **Discounting:** N/A  **Sensitivity analysis:** N/A | **Outcome/s of interest:** Mortality during a defined heatwave period.  **Types of costs measured:** No costs measured. | **Main finding:** Despite a sustained heatwave in England in 2013, the impact on mortality was considerably less than expected; a small cumulative excess of 195 deaths (95% confidence interval -87 to 477) in 65-year-olds and 106 deaths (95% CI -22 to 234) in 65-year olds was seen, nearly a fifth of excess deaths predicted based on observed temperatures. This impact was also less than seen in 2006 (2323 deaths) and 2003 (2234 deaths), despite a similarly prolonged period of high temperatures. The reasons for this are unclear and further work needs to be done to understand this and further clarify the predicted impact of increases in temperature.  **Additional finding:** Despite a prolonged period of increased temperatures, the impact of the 2013 heatwave on mortality was not large. The model used in this paper identifies cumulative mortality above the baseline during a defined heatwave period rather than directly attributing an increase to temperature; temporally associated factors other than temperature and environmental factors may be responsible for variation in mortality.  **Sensitivity analysis results (for economic evaluations):** No economic analysis conducted. |
| Hajat, S., Kovats, R. ., Atkinson, R. ., & Haines, A. (2002). Impact of hot temperatures on death in {London}: a time series approach. Journal of Epidemiology and Community Health, 56, 367. <https://www.ncbi.nlm.nih.gov/pmc/articles/PMC1732136/pdf/v056p00367.pdf>  (Hajat et al., 2002)  **Country:** England, UK  **Aim:** To investigate the relation between heat and mortality in London to determine the temperature threshold at which death rates increase and to quantify the effect of extreme temperatures on mortality. | **Intervention:** No intervention.  **Dates of data collection:**  Daily mortality counts in Greater London between January 1976 and December 1996.  **Population and sample size:**  186.54 deaths.  **Setting:** London, England.  **Delivery mode (e.g., remotely online, in person):** N/A  **Intervention deliverers:** N/A  **Timing and duration:** N/A  **Intervention description:** N/A | **Study type:** Quantitative.  **Length of follow-up:** N/A  **Type of economic evaluation/cost analysis:** No economic analysis.  **Perspective of analysis:** N/A  **Currency and cost year:** N/A  **Discounting:** N/A  **Sensitivity analysis:** N/A | **Outcome/s of interest:** Daily mortality counts in Greater London  **Types of costs measured:** No costs measured. | **Main finding:**  A plot of the basic mortality-temperature relation suggested that a rise in heat related deaths began at about 19°C. Average temperatures above the 97th centile value of 21.5°C (excluding those days from a 15 day “heatwave” period in 1976) resulted in an increase in deaths of 3.34% (95% CI 2.47% to 4.23%) for every one degree increase in average temperature above this value. It was found that the 1976 heatwave resulted in a particularly large number of deaths in comparison with other hot periods.  **Additional finding:**  The maximum daily death count of 467 occurred during a serious influenza epidemic (unrelated to heat) in February 1976.  There is a strong yearly seasonal pattern in the mortality series with most deaths occurring in the winter months of each year.  **Sensitivity analysis results (for economic evaluations):** No economic analysis conducted. |
| Heaviside, C., Vardoulakis, S., & Cai, X. M. (2016). Attribution of mortality to the urban heat island during heatwaves in the West Midlands, UK. Environmental Health: A Global Access Science Source, 15(Suppl 1). <https://doi.org/10.1186/s12940-016-0100-9>  (Heaviside et al., 2016)  **Country:** England, UK  **Aim:** To investigate the sensitivity of health impact estimates to the use of population weighting and the inclusion of urban temperatures in exposure data. | **Intervention:** No intervention.  **Dates of data collection:**  1993-2006.  **Population and sample size:** Population of the West Midlands, England in August 2003.  **Setting:** West Midlands, England.  **Delivery mode (e.g., remotely online, in person):** N/A  **Intervention deliverers:** N/A  **Timing and duration:** N/A  **Intervention description:** N/A | **Study type:** Quantitative (observational study).  **Length of follow-up:** No follow-up.  **Type of economic evaluation/cost analysis:** No economic analysis.  **Perspective of analysis:** N/A  **Currency and cost year:** N/A  **Discounting:** N/A  **Sensitivity analysis:** N/A | **Outcome/s of interest:** Mortality related to heatwave of 1^st^-10^th^ August 2003.  **Types of costs measured:** No costs measured. | **Main finding:**  The results suggest that the Urban Heat Island (UHI) contributed around 50 % of the total heat-related mortality during the 2003 heatwave in the West Midlands. We also find that taking a geographical, rather than population-weighted, mean of temperature across the regions under-estimates the population exposure to temperatures by around 1 °C, roughly equivalent to a 20 % underestimation in mortality.  **Additional finding:**  For a medium emissions scenario, a typical heatwave in 2080 could be responsible for an increase in mortality of around 3 times the rate in 2003 (278 vs. 90 deaths) when including changes in population, population weighting and the UHI effect in the West Midlands, and assuming no change in population adaptation to heat in future.  **Sensitivity analysis results (for economic evaluations):** No economic analysis conducted. |
| Johnson, H., Kovats, R. S., McGregor, G., Stedman, J., Gibbs, M., & Walton, H. (2005). The impact of the 2003 heat wave on daily mortality in England and Wales and the use of rapid weekly mortality estimates. *Euro Surveillance : Bulletin Européen Sur Les Maladies Transmissibles = European Communicable Disease Bulletin*, *10*(7), 168–171. <https://doi.org/10.2807/esm.10.07.00558-en>  (Johnson et al., 2005)  **Country:** England and Wales, UK  **Aim:** To get an estimate of the number of deaths attributable to the heat wave and reflected the pattern of daily deaths in relation to the hottest days. | **Intervention:** No intervention.  **Dates of data collection:** Mortality data were extracted from databases held by Office for National Statistics (ONS), for all deaths occurring on each day in July and August 2003, and for same months in the five preceding years, by age group (0–64, 65–74, 75 and over) and by Government Office Region (GOR).  **Population and sample size:** See above.  **Setting:** Regions in England and Wales.  **Delivery mode (e.g., remotely online, in person):** N/A  **Intervention deliverers:** N/A  **Timing and duration:** N/A  **Intervention description:** N/A | **Study type:** Quantitative. (observational study)  **Length of follow-up:** No follow-up.  **Type of economic evaluation/cost analysis:** No economic analysis.  **Perspective of analysis:** N/A  **Currency and cost year:** N/A  **Discounting:** N/A  **Sensitivity analysis:** N/A | **Outcome/s of interest:**  Mortality data  **Types of costs measured:** No costs measured. | **Main finding:**  The August 2003 heat wave was associated with a large short-term increase in mortality, particularly in London. Ozone and particulate matter concentrations were also elevated during the heat wave. Overall, there were 2139 (16%) excess deaths in England and Wales.  **Additional finding:**  The worst affected were people over the age of 75 years. The impact was greatest in the London region where deaths in those over the age of 75 increased by 59%. Estimated excess mortality was greater than for other recent heat waves in the United Kingdom.  **Sensitivity analysis results (for economic evaluations):** No economic analysis conducted. |
| Kovats, R. S., Hajat, S., & Wilkinson, P. (2004). Contrasting patterns of mortality and hospital admissions during hot weather and heat waves in Greater London, UK. Occupational and Environmental Medicine, 61(11), 893–898. <https://doi.org/10.1136/oem.2003.012047>  **Country:** England, UK  **Aim:** To model the effects of a heat using the average of the daily mean temperature over the index and previous two days.  To investigate the effects of hot weather and heat waves on emergency hospital admissions in Greater London, UK, for a range of causes and age groups. | **Intervention:** No intervention.  **Dates of data collection:** 1 April 1994 to 31 March 2000.  **Population and sample size: London.**  **Setting:** London, England, UK  **Delivery mode (e.g., remotely online, in person):** N/A  **Intervention deliverers:** N/A  **Timing and duration:** N/A  **Intervention description:** N/A | **Study type:** Quantitative (observational study).  **Length of follow-up:** No follow-up.  **Type of economic evaluation/cost analysis:** No economic analysis.  **Perspective of analysis:** N/A  **Currency and cost year:** N/A  **Discounting:** N/A  **Sensitivity analysis:** N/A | **Outcome/s of interest:** Hospital admissions in a heatwave in London, UK.  **Types of costs measured:** No costs measured. | **Main finding:**  There was no clear evidence of a relation between total emergency hospital admissions and high ambient temperatures, although there was evidence for heat related increases in emergency admissions for respiratory and renal disease, in children under 5, and for respiratory disease in the 75+ age group.  **Additional finding:**  During the heat wave of 29 July to 3 August 1995, hospital admissions showed a small non-significant increase: 2.6% (95% CI 22.2 to 7.6), while daily mortality rose by 10.8% (95% CI 2.8 to 19.3) after adjusting for time varying confounders.  **Sensitivity analysis results (for economic evaluations):** No economic analysis conducted. |
| Leonardi, G. S., Hajat, S., Kovats, R. S., Smith, G. E., Cooper, D., & Gerard, E. (2006). Syndromic surveillance use to detect the early effects of heat-waves: An analysis of NHS direct data in England. Sozial- Und Praventivmedizin, 51(4), 194–201. [**https://doi.org/10.1007/s00038-006-5039-0**](https://doi.org/10.1007/s00038-006-5039-0)  (Leonardi et al., 2006)  **Country:** England, UK  **Aim:** To investigate the effects of high ambient temperatures, including the summer 2003 heat-episode, on NHS Direct usage and its suitability as a surveillance tool in heat health warning systems | **Intervention:** No intervention.  **Dates of data collection:**  NHS Direct call data from Dec 2001 to May 2004. Daily meteorological data was also obtained.  **Population and sample size:**  Analysis  **Setting:** NHS direct data, focussing on the south of England.  **Delivery mode (e.g., remotely online, in person):** -No intervention. Syndromic surveillance  **Intervention deliverers:** No intervention.  **Timing and duration:**  Cross section.  **Intervention description:** No intervention. | **Study type:** Quantitative. Syndromic surveillance. Time series methods used to investigate associations between temp and NHS Direct calls.  **Length of follow-up:** No follow-up.  **Type of economic evaluation/cost analysis:** No economic analysis.  **Perspective of analysis:** N/A  **Currency and cost year:** N/A  **Discounting:** N/A  **Sensitivity analysis:** N/A | **Outcome/s of interest:**  Daily rates of all symptomatic calls, and daily proportion of calls for selected causes (fever, vomiting, difficulty breathing, heat-/sunstroke)  **Types of costs measured:** No costs measured. | **Main finding:**  Total calls were moderately increased as environmental temperature increased; this effect was greatest in calls for young children and for fever. Increase in fever calls per 10 degree temp increase was significant for infants 0-4 P <0.001), and older adults 65+ (P = 0.001). Total calls were moderately elevated during two summer heat episodes in 2003: calls specifically for heat/sun stroke increased acutely in response to these episodes. No association was apparent between environmental temperature and proportion of calls for vomiting and difficulty breathing.  **Additional finding:**  Heat wave episodes investigated separately but lacked power to create robust models. Increase in calls for heat stroke.  The apparent moderate effect of high temps on morbidity in the elderly requires further explanation. They feel that the results may underrepresent their morbidity rather than constitute evidence of an absence of a major heat effect, as the mortality rate is known to rise in heatwaves. It seems many that die in heatwaves do not access healthcare. Older people may have poorer perception of ambient temperature.  More research needed on mechanisms by which heat related morbidity occurs.  **Sensitivity analysis results (for economic evaluations):**  **Recommendations:** No economic analysis conducted. |
| Oven, K. J., Curtis, S. E., Reaney, S., Riva, M., Stewart, M. G., Ohlemüller, R., Dunn, C. E., Nodwell, S., Dominelli, L., & Holden, R. (2012). Climate change and health and social care: Defining future hazard, vulnerability and risk for infrastructure systems supporting older people’s health care in England. Applied Geography, 33(1), 16–24. <https://doi.org/10.1016/j.apgeog.2011.05.012>  (Oven et al., 2012)  **Country:**  England, UK  **Aim:** To identify areas of England where changing environmental conditions and demographic trends are likely to make it particularly urgent to assess the vulnerability of infrastructure and develop adaptation and resilience strategies for older people’s care during extreme weather-related events in the future. | **Intervention:** No intervention.  **Dates of data collection:**  The authors considered the latest climate projections for the 2030s from the UK Climate Impacts Programme (UKCP09); river and coastal flooding projections for the 2050s from the 2004 UK Government’s Foresight Flood and Coastal Defence Project (Environment Agency, 2004); and demographic projections for 2031 produced by the Office for National Statistics, UK.  **Population and sample size:** See above  **Setting:** England  **Delivery mode (e.g., remotely online, in person):** N/A  **Intervention deliverers:** N/A  **Timing and duration:** N/A  **Intervention description:** N/A | **Study type:** Quantitative (Modelling/Mapping study).  **Type of economic evaluation/cost analysis:** No economic analysis.  **Perspective of analysis:** N/A  **Currency and cost year:** N/A  **Discounting:** N/A  **Sensitivity analysis:** N/A | **Outcome/s of interest:** Mapping exercises, like the one summarised reflect this degree of complexity and may be used as tools for local resilience planning.  **Types of costs measured:** No costs measured. | **Main finding:** Risk to build infrastructure supporting older people’s care should be assessed in terms of multiple facets of hazard and vulnerability. This will be quite challenging for local agencies with limited resources attempting to create resilient environments and people.  Additional findings: Outputs from the Foresight project (Environment Agency, 2004) suggest the most significant change in the annual probability of flooding will occur in the South East of England, the East of England and the Yorkshire and Humber region (Fig. 2i). These areas (shown in dark grey) will experience an increase in the annual probability (frequency) of inundation by as much as 4135% and are areas where flood response strategies are most likely to need to be adjusted and enhanced in future. The areas susceptible to the greatest projected increase in flooding are either coastal or located along major estuaries, and are affected by subsidence and sea-level rise, rather than fluvial flooding.  **Sensitivity analysis results (for economic evaluations):**  No economic analysis conducted. |
| Page LA, Hajat S, Kovats RS, Howard LM. Temperature-related deaths in people with psychosis, dementia and substance misuse. Br J Psychiatry. 2012 Jun;200(6):485-90. doi: 10.1192/bjp.bp.111.100404.  (Page et al., 2012)  **Country:** UK  **Aim:**  To estimate risk conferred by high ambient temperature on patients with psychosis, dementia and substance misuse. | **Intervention:**  N/A  **Dates of data collection:**  1 January 1998 and 31 December 2007  **Intervention recipients and sample size:**  N/A  **Setting:**  England  **Delivery mode (e.g., remotely online, in person):**  N/A  **Intervention deliverers:**  N/A  **Timing and duration:**  N/A  **Intervention description:**  N/A | **Study type:**  Observational study  **Length of follow-up:**  No follow-up period  **Type of economic evaluation/cost analysis**  N/A  **Perspective of analysis:**  Applying time-series regression analysis to assess the short-term relationship between mortality level and daily fluctuations in temperature.  **Currency and cost year:**  N/A  **Discounting:**  N/A  **Sensitivity analysis:**  N/A | **Outcome/s of interest:**  Mortality level  **Types of costs measured:**  N/A | **Main finding:**  Patients with mental illness showed an overall increase in risk of death of 4.9% (95% CI 2.0–7.8) per 1℃ increase in temperature above the 93rd percentile of the annual temperature distribution. Younger patients and those with a primary diagnosis of substance misuse demonstrated the greatest mortality risk.  The increased risk of death during hot weather in patients with psychosis, dementia and substance misuse has implications for public health strategies during heatwaves.  **Additional finding:**  N/A  **Sensitivity analysis results (for economic evaluations):**  N/A |
| Page LA, Hajat S, Kovats RS. Relationship between daily suicide counts and temperature in England and Wales. British Journal of Psychiatry. 2007;191(2):106-112. doi:10.1192/bjp.bp.106.031948  (Page et al., 2007)  **Country:**  UK  **Aim:**  To assess the relationship  between daily temperature and daily  suicide counts in England and Wales  between 1 January 1993 and 31 December  2003 and to establish whether heatwaves  are associated with increased mortality  from suicide. | **Intervention:**  N/A  **Dates of data collection:**  between 1 January 1993 and 31 December 2003  **Intervention recipients and sample size:**  N/A  **Setting:**  England, Wales  **Delivery mode (e.g., remotely online, in person):**  N/A  **Intervention deliverers:**  N/A  **Timing and duration:**  N/A  **Intervention description:**  N/A | **Study type:**  Quantitative study  **Length of follow-up:**  No follow-up period  **Type of economic evaluation/cost analysis**  N/A  **Perspective of analysis:**  Time-series regression  analysis (using  Poisson generalized linear modelling) was used to explore and quantify  the relationship between daily suicide  counts and daily temperature.  **Currency and cost year:**  N/A  **Discounting:**  N/A  **Sensitivity analysis:**  N/A | **Outcome/s of interest:**  The relationship between daily suicide  counts and daily temperature.  **Types of costs measured:**  N/A | **Main finding:**  No spring or summer peak in suicide was found. Above 18 ℃, each 1℃ increases in mean temperature was associated with a 3.8 and 5.0% rise in  suicide and violent suicide respectively.  Suicide increased by 46.9% during the  1995 heatwave, whereas no change was seen during the 2003 heatwave.  There is increased risk of suicide during hot weather.  **Additional finding:**  N/A  **Sensitivity analysis results (for economic evaluations):**  N/A |
| Rendell, Rebecca & Higlett, Michael & Khazova, Marina & O'Hagan, John. (2020). Public Health Implications of Solar UV Exposure during Extreme Cold and Hot Weather Episodes in 2018 in Chilton, South East England. Journal of Environmental and Public Health. 2020. 1-9. 10.1155/2020/2589601.  (Rendell et al., 2020)  **Country:** England, UK  **Aim:**  To assess public health impacts of solar UV exposure, both positive and negative, during extreme hot and cold weather in England in 2018. | **Intervention:**  Not an intervention study  **Dates of data collection:**  2018  **Intervention recipients and sample size:**  N/A  **Setting:**  Chilton, South East England.  **Delivery mode (e.g., remotely online, in person):**  N/A  **Intervention deliverers:**  N/A  **Timing and duration:**  N/A  **Intervention description:**  N/A | **Study type:** Quantitative study  **Length of follow-up:**  N/A  **Type of economic evaluation/cost analysis**  N/A  **Perspective of analysis:**  The implications of cold and hot weather periods for public health in terms of sun exposure are assessed regarding the extreme temperatures experienced and people’s likely behaviour, the current UK alert system and public health advice.  **Currency and cost year:**  N/A  **Discounting:**  N/A  **Sensitivity analysis:**  N/A | **Outcome/s of interest:**   - Extreme temperature experience - People’s likely behaviour - The current alert system - The public health advice   **Types of costs measured:**  N/A | **Main finding:**  No key findings related to health impact of heat/high temperature/UV daily doses  **Additional finding:**  1) The public health  implications are complex and highly dependent on behaviour and sociodemographic variables such as skin colour.  2) For all three  periods, significant positive correlation between maximum daily temperature and erythema-effective UV daily dose were found.  3)Public health advice may be improved by taking account of both temperature  and UV and their implications for behaviour**.**  **Sensitivity analysis results (for economic evaluations):**  N/A |
| Rizmie, D., de Preux, L., Miraldo, M., & Atun, R. (2022). Impact of extreme temperatures on emergency hospital admissions by age and socio-economic deprivation in England. Social Science and Medicine, 308(July), 115193. <https://doi.org/10.1016/j.socscimed.2022.115193>  (Rizmie et al., 2022)  **Country:** England, UK  **Aim:** To estimate the impact of extreme temperatures on emergency hospital admissions by age and socio-economic deprivation in England. | **Intervention:** No intervention.  **Dates of data collection:** The authors examined the time period between 2001 and 2012  **Population and sample size:** Population of England.  **Setting:** Healthcare setting, England.  **Delivery mode (e.g., remotely online, in person):** N/A  **Intervention deliverers:** N/A  **Timing and duration:** N/A  **Intervention description:** N/A | **Study type:** Quantitative (modelling study).  **Length of follow-up:** No follow-up.  **Type of economic evaluation/cost analysis:** No economic analysis.  **Perspective of analysis:** N/A  **Currency and cost year:** N/A  **Discounting:** N/A  **Sensitivity analysis:** N/A | **Outcome/s of interest:** Emergency hospital admissions across the entire population of England.  **Types of costs measured:** No costs measured. | **Main finding:**  Larger hospitalisation impacts were associated with extreme cold temperatures than with extreme hot temperatures. The less extreme temperatures produce admission patterns like their extreme counterparts, but at lower magnitudes. Results also showed an increase in admissions with extreme temperatures that were more prominent among older and socioeconomically deprived populations, particularly across admissions for metabolic diseases and injuries.  The effect of extreme heat, in comparison to 10–15◦C, was positive  and statistically significant for admissions across all disease categories, except from circulatory diseases and neoplasms. This increase in ad- missions was smaller for respiratory diseases (9.2%) and injuries (6.0%). Metabolic diseases had the largest effect magnitude, suggesting that, at temperatures above 30◦C, there was an increase of 25.8% in admissions, per day per hospital, relative to a day between 10◦C and 15◦C. This was followed by an increase of 21.1% admissions for infectious diseases (p < 0.001).  **Additional finding:** Our findings show that hospitals experience an increase in admissions with both extreme hot and cold temperatures. The less extreme temperatures produce admission patterns like their extreme counter- parts, but at lower magnitudes.  **Sensitivity analysis results (for economic evaluations):**  No economic analysis conducted. |
| Rooney, C., McMichael, A. J., Kovats, R. S., & Coleman, M. P. (1998). Excess mortality in England and Wales, and in Greater London, during the 1995 heatwave. Journal of Epidemiology and Community Health, 52(8), 482–486. <https://doi.org/10.1136/jech.52.8.482>  (Rooney et al., 1998)  **Country:** England, UK  **Aim:** To assess the impact on mortality of the heatwave in England and Wales during July and August 1995 and to describe any difference in mortality impact between the Greater London urban population and the national population. | **Intervention:** No intervention.  Analysis of variation in daily mortality in England and Wales and in Greater London during a five-day heatwave in July and August 1995, by age, sex, and cause.  **Dates of data collection:**  1995.  **Population and sample size:**  Mortality statistics from Office for National Statistics. CET (Central England Temperature) data and London data obtained from UK Meteorological Office.  **Setting:** England, Wales and Greater London.  **Delivery mode (e.g., remotely online, in person):** -No intervention.  **Intervention deliverers:** No intervention.  **Timing and duration:**  Cross section.  **Intervention description:** No intervention. | **Study type:** Quantitative. Analysis of mortality data and air pollution mortality risk coefficients.  **Length of follow-up:** No follow-up.  **Type of economic evaluation/cost analysis:** No economic analysis.  **Perspective of analysis:** N/A  **Currency and cost year:** N/A  **Discounting:** N/A  **Sensitivity analysis:** N/A | **Outcome/s of interest:**  Mortality increases during a heatwave.  Association of mortality with air pollution risk.  Comparison of London vs England/Wales as a whole.  **Types of costs measured:** No costs measured. | **Main finding:**  Exceptionally high temperatures in England and Wales do cause increases in daily mortality.  An estimated 619 extra deaths (8.9% increase, approximate 95% confidence interval 6.4, 11.3%) were observed during this heatwave in England and Wales.  Excess deaths were apparent in all age groups, most noticeably in women and for deaths from respiratory (12.4%) and cerebrovascular disease (11.3%).  Using published daily mortality risk coefficients for air pollutants in London, it was estimated that up to 62% of the excess mortality in England and Wales during the heatwave may be attributable to concurrent increases in air pollution in London, and 38% in the rest of England/Wales.  **Additional finding:**  Higher nighttime temperatures in London and greater deprivation may contribute to excess mortality there.  **Sensitivity analysis results (for economic evaluations):**  **Recommendations:** No economic analysis conducted. |
| Sahani , J., Kumar, P., Debele, S., & Emmanuel, R. (2022). Heat risk of mortality in two different regions of the United Kingdom. Sustainable Cities and Society, 80(February), 103758. <https://doi.org/10.1016/j.scs.2022.103758>  (Sahani et al., 2022)  **Country:** England and Scotland, UK  **Aim:** This study aims to associate temperature and mortality and explore its temporal variation. | **Intervention:** No intervention.  **Dates of data collection:**  2014-2015.  **Population and sample size:**  Used data from 1981 to 2018 – 38 years.  **Setting:** Southeast England and Aberdeenshire, Scotland.  **Delivery mode (e.g., remotely online, in person):**  No intervention.  **Intervention deliverers:** No intervention.  **Timing and duration:** N/A  **Intervention description:** No intervention. | **Study type:** Quantitative. A Poisson regression model combined with a distributed lag non-linear model was applied over daily mortality and maximum temperature data from 1981 to 2018 to formulate the lagged response of summer temperature. The relative risk (RR) and mortality attributable fraction with respect to minimum mortality temperature (MMT) in Southeast England and Aberdeenshire, Scotland, UK was calculated  **Length of follow-up:** No follow-up.  **Type of economic evaluation/cost analysis:** No economic analysis.  **Perspective of analysis:** N/A  **Currency and cost year:** N/A  **Discounting:** N/A  **Sensitivity analysis:** N/A | **Outcome/s of interest:**  Relative Risk and Mortality Attributable Fraction.  **Types of costs measured:** No costs measured. | **Main finding:** Relative risk (RR) is higher in the 2000–2018 sub-period than in 1981–1999 in SE England. Overall, RR for very high temperatures within a lag of around 2–3 days is quite high and increases exponentially. The risk, however, reduces and becomes nearly constant around a lag of 5 or more days. • The RR has reduced in the second sub-period in Aberdeenshire but showed a varying pattern of lagged effect for very high temperatures indicating a stronger harvesting effect than in SE England. In Southeast England, the RR for high and extreme (95th and 99th percentile) temperatures are 2 and 7 percent higher than at MMT respectively. In Aberdeenshire, these are negligible and excess 4% respectively.  **Additional finding:**  The risk for high and extreme temperature has increased by 1% and 7% in SE England, but it has decreased by 2% and 6% in Aberdeenshire considering two 10-year sub-periods. Similarly, the AF has increased for SE England but has decreased for Aberdeenshire. The increases are 1.11, 1.33 and 1.9 times in SE England and the decreases are 0.51, 0.49 and 0.15 times for hot, 95th and 99th percentile temperatures respectively considering the two sub-periods.  **Sensitivity analysis results (for economic evaluations):** No economic analysis conducted.  **Recommendations:** Need to consider housing conditions, socioeconomic conditions, segregating mortality type (by disease/age/sociodemographic factors) to calculate risk for particular section of vulnerable population, which can help formulate a more focused heat mitigation plan. |
| Smith, S., Elliot, A. J., Hajat, S., Bone, A., Bates, C., Smith, G. E., & Kovats, S. (2016). The impact of heatwaves on community morbidity and healthcare usage: A retrospective observational study using real-time syndromic surveillance. International Journal of Environmental Research and Public Health, 13(1). <https://doi.org/10.3390/ijerph13010132>  (Smith, Elliot, Hajat, Bone, Bates, et al., 2016)  DOI: doi:10.3390/ijerph13010132  **Country:** England, UK  **Aim:** the numbers of heatstroke cases are proportionally low. Therefore, the aim of this work was to consider the potential sensitivity of more common morbidity outcomes to inform the planning and response to heatwaves in England.  A second aim was to examine a subset of high-risk individuals to see if they were more likely to indicate an increase in heat-related morbidity. | **Intervention:** No intervention. Investigated the impact of a moderate heatwave on a range of presenting comorbidities in England  **Dates of data collection:**  2014-2015.  **Population and sample size:**  Data stratified by age group and compared between a heatwave year (2013) and non-heatwave years (2012 and 2014).  Incidence ratios used to estimate differential impact of heatwave compared to non-heatwave summers.  **Setting:** England  **Delivery mode (e.g., remotely online, in person):** -No intervention.  **Intervention deliverers:** No intervention.  **Timing and duration:** N/A  **Intervention description:** No intervention. | **Study type:** Quantitative. The authors used syndromic surveillance to achieve the aims as the systems can provide real-time intelligence during a heatwave, a comprehensive picture of national activity, historic data can be used to identify current anomalies and they can be used to monitor a range of morbidity indicators in the general population.  **Length of follow-up:** No follow-up.  **Type of economic evaluation/cost analysis:** No economic analysis.  **Perspective of analysis:** N/A  **Currency and cost year:** N/A  **Discounting:** N/A  **Sensitivity analysis:** N/A | **Outcome/s of interest:**  Asthma, breathing difficulty, cerebrovascular accident and cardiovascular symptoms.  **Types of costs measured:** No costs measured.  Limitations: the two non-heatwave years did contain a small number of level 2 alerts. | **Main finding:** There were no apparent differences for the morbidities tested between the 2013 heatwave and non-heatwave years. A subset of GPIH data were used to study individuals at higher risk from heatwaves based on their pre-existing disease.  The results of this current study illustrate that syndromic surveillance systems monitoring signs and symptoms of respiratory complaints such as asthma, wheeze and difficulty breathing were not sensitive to periods of hot weather. In addition, there was no evidence of increased incidence of CVA or myocardial infarction/ischaemia during the 2013 heatwave period compared to other years. This is consistent with other studies using hospital admissions data which has shown the myocardial infarction risk increase with cold but not with heat.  **Additional finding:**  Higher risk patients were not more likely to present at GPs or ED than other individuals. Comparing GPIH consultations and ED attendances for myocardial infarction/ischaemia, there was evidence of a fall in the presentation of myocardial infarction during the heatwave, which was particularly noted in the 65–74 years age group (and over 75 years in ED attendances).  **Sensitivity analysis results (for economic evaluations):** No economic analysis conducted.  **Recommendations:** The decrease in myocardial ischaemia ED attendances reported requires further work to elucidate the underlying drivers for this fall.  The results from this work suggest that there is no additional benefit in monitoring a range of health morbidity outcomes in addition to heatstroke symptoms within the England heatwave plan surveillance system. Previous work has demonstrated that monitoring the presentation of heatstroke symptoms provides a good indicator of the impact of heat across different health systems in the community [5], and in conjunction with these findings this will continue to be the main outcome measure and input to the heatwave plan. Monitoring mortality rates during heatwaves still represents the most effective measure of the severe impact of heat and systems are in place within England to monitor all-cause mortality during heatwaves in near real-time. |
| Smith, S., Elliot, A. J., Hajat, S., Bone, A., Smith, G. E., & Kovats, S. (2016). Estimating the burden of heat illness in England during the 2013 summer heatwave using syndromic surveillance. Journal of Epidemiology and Community Health, 70(5), 459–465. https://doi.org/10.1136/jech-2015-206079  (Smith, Elliot, Hajat, Bone, Smith, et al., 2016)  DOI: doi:10.1136/jech-2015-206079  **Country:** England, UK  **Aim:** The objective of this paper was to describe the impact of the 2013 heatwave on morbidity utilising syndromic surveillance systems, using reported cases of heat illness presenting to healthcare services in England. | **Intervention:** No intervention.  **Dates of data collection:** 3 – 23^rd^ July 2013 (heatwave).  **Population and sample size:**  General practitioner in hours (GPIH), GP out of hours (GPOOH) and ED syndromic surveillance systems were used to monitor the health impact of heat/sun stroke symptoms (heat illness). Data were stratified by age group and compared between heatwave and non-heatwave years. Incidence rate ratios were calculated for GPIH heat illness consultations.  **Setting:** Nationwide  **Delivery mode (e.g., remotely online, in person):** -No intervention.  **Intervention deliverers:** No intervention.  **Timing and duration:**  3 – 23^rd^ July 2013 (heatwave)  **Intervention description:** No intervention. | **Study type:** Quantitative. Syndromic surveillance.  **Length of follow-up:** No follow-up.  **Type of economic evaluation/cost analysis:** No economic analysis.  **Perspective of analysis:** N/A  **Currency and cost year:** N/A  **Discounting:** N/A  **Sensitivity analysis:** N/A | **Outcome/s of interest:**  Attendance at GP in hours, GP OOH and ED for heat stroke related symptoms.  **Types of costs measured:** No costs measured.  Limitations:  Used the area covered by GPIH system to estimate total heat illness for England, assuming all areas of the country were equally affected by hot weather. | **Main finding:**  GP consultations and ED attendances for heat illness increased during the heatwave period; GPIH consultations increased across all age groups, but the highest rates were in school children and those aged ≥75 years, with the latter persisting beyond the end of the heatwave. Extrapolating to the English population, the authors estimated that the number of GPIH consultations for heat illness during the whole summer (May to September) 2013 was 1166 (95% CI 1064 to 1268). This was double the rate observed during non-heatwave years.  **Additional finding:**  There is a lag following a heatwave – illnesses in the >75 group continued into early August.  **Sensitivity analysis results (for economic evaluations):**  No economic analysis conducted.  **Recommendations:**  Findings support the monitoring of heat illness as part of the Heatwave Plan for England. |
| Thompson R, Landeg O, Kar-Purkayastha I, Hajat S, Kovats S, O'Connell E. Heatwave Mortality in Summer 2020 in England: An Observational Study. Int J Environ Res Public Health. 2022 May 18;19(10):6123. doi: 10.3390/ijerph19106123.  (Thompson et al., 2022)  **Country:**  England, UK  **Aim:**  To gain insight into how the impact of the 2020 heatwaves was different  from other years, in terms of cause of death and place of death, and if these differences can be attributed to the COVID-19 pandemic and related countermeasures. | **Intervention:**  Not an intervention study  **Dates of data collection:**  01 June to 15 September 2020 (death record from the Office for National Statistics (ONS))  **Intervention recipients and sample size:**  N/A  **Setting:**  England  **Delivery mode (e.g., remotely online, in person):**  N/A  **Intervention deliverers:**  N/A  **Timing and duration:**  N/A  **Intervention description:**  N/A | **Study type:**  Observational study  **Length of follow-up:**  No follow-up period  **Type of economic evaluation/cost analysis**  N/A  **Perspective of analysis:**  Episode analysis; The total and cause-specific mortality in  2020 was compared to previous heatwave events in England.  **Currency and cost year:**  N/A  **Discounting:**  N/A  **Sensitivity analysis:**  N/A | **Outcome/s of interest:**  Heatwave Impacts by Age, Cause of Death, Place of Death and Region  **Types of costs measured:**  N/A | **Main finding:**  The impact of the heatwave in 2020 was significantly larger than for previous events.  The main causes of death which observed a significant increase in 2020 heatwaves included cardiovascular disease, respiratory disease, and Alzheimer’s disease and Dementia  **Additional finding:**  There was no evidence that the 2020 heatwaves increased the proportion of people dying in different settings, or that the pandemic shifted the main underlying causes of death during a heatwave.  **Sensitivity analysis results (for economic evaluations):**  N/A |
| Wan, K., Feng, Z., Hajat, S., & Doherty, R. M. (2022). Temperature-related mortality and associated vulnerabilities: evidence from Scotland using extended time-series datasets. Environmental Health: A Global Access Science Source, 21(1), 1–14. https://doi.org/10.1186/s12940-022-00912-5  (Wan et al., 2022)  **Country:** Scotland, UK  **Aim:** Aims to investigate the effects of extremes of temperature. The three main objectives of this study are to a) investigate the relationship between ambient daily temperature and mortality risk; b) explore trends in the cold/heat related mortality risk over time; c) assess the variation of cold/heat-related mortality risk by demographic and socio-economic features including age-group, sex, marital status, area-level deprivation and cause of death.  Suggested that Scottish population, which is understudied relative to England, may be more vulnerable to temperature extremes due to the already higher mortality rate (inequality, negative health behaviours, deprivation).  NB. England has had a heatwave plan for 20 years, whilst the devolved nations, including Scotland, do not. | **Intervention:** No intervention.  Uses time-series regression analysis with distributed lag non-linear models to characterise acute relationships between daily mean ambient temperature and mortality in Scotland.  **Dates of data collection:** 1974 - 2018  **Population and sample size:**  Scottish population  **Setting:** the four largest cities (Aberdeen, Dundee, Edinburgh and Glasgow) and three regions - northern, western and eastern Scotland  **Delivery mode (e.g., remotely online, in person):** -No intervention.  **Intervention deliverers:** No intervention.  **Timing and duration:** N/A  **Intervention description:** No intervention.  Daily mortality counts in Scotland between 1974–2018 were obtained from National Records of Scotland.  Daily maximum and minimum ambient temperatures were obtained from HadUK-Grid Gridded Climate Observation and downloaded from the Centre for Environmental Data Analysis [6. The daily mean temperature of each locality was taken as the average of the area-level mean daily maximum and minimum temperature.  Due to data limitations, only data on relative humidity, particulate matter with an aerodynamic diameter smaller than 10 µm (PM10), and ozone for Edinburgh during 2004–2018 were available for sensitivity analysis. | **Study type:** Quantitative.  Increases in mortality risk under extreme cold and heat in individual cities and regions were aggregated using multivariate meta-analysis. Cold results are summarised by comparing the relative risk (RR) of death at the 1st percentile of localised temperature distributions compared to the 10th percentile, and heat effects as the RR at the 99th compared to the 90th percentile.  **Length of follow-up:** No follow-up.  **Type of economic evaluation/cost analysis:** No economic analysis.  **Perspective of analysis:** N/A  **Currency and cost year:** N/A  **Discounting:** N/A  **Sensitivity analysis:** N/A | **Outcome/s of interest:**  Temperature-related mortality and associated vulnerabilities.  **Types of costs measured:** No costs measured. | **Main finding:** Adverse cold effects were observed in all cities and regions, and heat effects were apparent in all cities and regions. Aggregate all-cause mortality risk in Scotland was estimated to increase by 9% (95% confidence interval, CI: 8%, 11%) under extreme cold and 4% (CI: 3%, 5%) under extreme heat.  **Additional finding:** The elderly had the highest RR under both extreme cold and heat. Males experienced greater cold effects than females, whereas the reverse was true with heat effects, particularly among the elderly. Those who were unmarried had higher RR than those married under extreme heat, and the effect remained after controlling for age. The younger population living in the most deprived areas experienced higher cold and heat effects than in less deprived areas. Deaths from respiratory diseases were most sensitive to both cold and heat exposures, although mortality risk for cardiovascular diseases was also heightened, particularly in the elderly. Cold effects were lower in the most recent 15 years, which may be linked to policies and actions in preventing the vulnerable population from cold impacts. No temporal trend was found with the heat effect.  Mortality risk started increasing as the temperatures increase above around 14.5C in Scotland. This indicates that the heat-health impacts can also be observed at relatively low temperatures in places with a cool climate.  **Sensitivity analysis results (for economic evaluations):** N/A  **Recommendations:** The study identifies those groups who would benefit most from targeted actions to reduce cold- and heat-related mortalities and suggests further research to identify modifiable factors in these groups, and to consider future health burdens under climate change scenarios. This research is needed to inform future policy in Scotland. |
| Zhang S, Breitner S, Rai M, Nikolaou N, Stafoggia M, De' Donato F, Samoli E, Zafeiratou S, Katsouyanni K, Rao S, Palomares AD, Gasparrini A, Masselot P, Aunan K, Peters A, Schneider A. Assessment of short-term heat effects on cardiovascular mortality and vulnerability factors using small area data in Europe. Environ Int. 2023 Sep;179:108154. doi: 10.1016/j.envint.2023.108154. Epub 2023 Aug 16.  (Zhang et al., 2023)  **Country:** UK  **Aim:**  To assess heat effects on cardiovascular disease (CVD) mortality and potential vulnerability factors using data from three European countries, including urban and rural settings. | **Intervention:**  Not an intervention study  **Dates of data collection:**  May-September 2000 – 2018 (CVD deaths record)  **Intervention recipients and sample size:**  N/A  **Setting:**  England and Wales  **Delivery mode (e.g., remotely online, in person):**  N/A  **Intervention deliverers:**  N/A  **Timing and duration:**  N/A  **Intervention description:**  N/A | **Study type:**  Observational study  **Length of follow-up:**  No follow-up period.  **Type of economic evaluation/cost analysis**  N/A  **Perspective of analysis:**  Applied area-specific Quasi-Poisson regression using  distributed lag nonlinear models to examine the heat effects on CVD mortality  **Currency and cost year:**  N/A  **Discounting:**  N/A  **Sensitivity analysis:**  Sensitive analysis using minimum mortality temperature (MMT) as the reference point. | **Outcome/s of interest:**  Short-term association between heat and CVD mortality  **Types of costs measured:**  N/A | **Main finding:**  When pooling the heat effects across three countries, the author observed a  heat-related increase in the risk for CVD mortality during the warm season.  Greater heat vulnerability was observed in areas with high population density, high degree of urbanization, low green coverage, and high levels of fine particulate matter.  **Additional finding:**  Women were more affected by heat.  **Sensitivity analysis results (for economic evaluations):**  N/A |

**END**
