## Supplementary File 4 for "Extreme weather events in the UK and resulting public health outcomes"

### **Supplementary file 4: Extreme wind study**

| Goldman, A., Eggen, B., Golding, B., & Murray, V. (2014). The health impacts of windstorms: A systematic literature review. Public Health, 128(1), 3–28. <https://doi.org/10.1016/j.puhe.2013.09.022>  (Goldman et al., 2014)  **Country:** England, UK  **Aim: To** review the international evidence on the impacts of windstorms on human health. | **Intervention:** No intervention.  **Dates of data collection:**  Searches started from 1946 to May 2012.  **Population and sample size:**  A total of 69 papers were included in this review.  **Setting:** Worldwide  **Delivery mode (e.g., remotely online, in person):** -No intervention.  **Intervention deliverers:** N/A.  **Timing and duration:** N/A  **Intervention description:** N/A. | **Study type:** A review of international literature.  **Length of follow-up:** No follow-up.  **Type of economic evaluation/cost analysis:** No economic analysis.  **Perspective of analysis:** N/A  **Currency and cost year:** N/A  **Discounting:** N/A  **Sensitivity analysis:** N/A | **Outcome/s of interest:** The health effects of high-velocity windstorms.  **Types of costs measured:** No costs measured. | **Main finding:**  Human health can be severely affected by windstorms. Direct effects occur during the impact phase of a storm, causing death and injury due to the force of the wind. Becoming airborne, being struck by flying debris or falling trees and road traffic accidents are the main dangers. Indirect effects, occurring during the pre- and post-impact phases of the storm, include falls, lacerations, and puncture wounds, and occur when preparing for, or cleaning up after a storm. Power outages are a key issue and can lead to electrocution, fires and burns and carbon monoxide poisoning from gasoline powered electrical generators.  **Additional finding:**  Additionally, worsening of chronic illnesses due to lack of access to medical care or medication can occur. Other health impacts include infections and insect bites.  Only two peer reviewed reports on the health impacts of windstorms in the UK were identified in this review.  **Sensitivity analysis results (for economic evaluations):**  **Recommendations:** No economic analysis conducted. |
| --- | --- | --- | --- | --- |

END
