## Supplementary File 5 for "Extreme weather events in the UK and resulting public health outcomes"

### **Supplementary file 5 – Table of abbreviations**

| **Abbreviation** | **Description** |
| --- | --- |
| AOR | Adjusted Odds Ratio |
| CI | Confidence Interval |
| CMD | Common Mental Disorder |
| CMH | Common Mental Health |
| eCI | Empirical Confidence Interval |
| EWE | Extreme Weather Event |
| EWEs | Extreme Weather Events |
| GP | General Practitioner |
| GPOOC | General Practitioner Out of Hours Consultations |
| MMT | Minimum Mortality Temperature |
| OECD | Organisation for Economic Co-operation and Development |
| PTSD | Post-Traumatic Stress Disorder |
| NHS | National Health Service |
| RR | Relative Risk |
| SR | Systematic Review |
| UHI | Urban Heat Island |
| UV | Ultraviolet |
| USA | United States of America |
| UK | United Kingdom |
