## Supplementary File 6 for "Extreme weather events in the UK and resulting public health outcomes"

### Supplementary file 6 - Quality appraisal tables

As the quality appraisal tools varied in the number of questions that they asked, the research team decided on the following system for reaching a quality rating:

**11 checkboxes** - Cut off scores are defined in this review as; 11 to 9 out of 11 – high quality, 6 to 8 out of 11 – moderate quality, 0 to 5 out of 11 – low quality.

**10 checkboxes** – Cut off scores could be 10 to 8 out of 10 – high quality, 7 to 5 out of 10 - moderate quality, 0 to 4 out of 10 – low quality.

**8 checkboxes** – Cut off scores could be 7 to 8 out of 8 – high quality, 5 to 6 out of 8 – moderate quality, 0 to 4 out of 8 – low quality.

#### Flooding studies: Quality appraisal

For the flooding papers, the Joanna Briggs quality appraisal tools for cross-sectional studies, surveys and for qualitative studies were used (Joanna Briggs Institute, 2017b, 2017a). See Tables below.

##### Table 6.1 JBI critical appraisal checklist for systematic reviews and research synthesis (Joanna Briggs Institute, 2017a)

| **Study** | **JBI Appraisal items** | | | | | | | | | | | **Score** |
| --- | --- | --- | --- | --- | --- | --- | --- | --- | --- | --- | --- | --- |
|  | **1** | **2** | **3** | **4** | **5** | **6** | **7** | **8** | **9** | **10** | **11** |  |
| Euripidou and Murray (2004) | Y | N | N | N | N | N | N | N | N | N | Y | 2/11 Low |
| Hunter (2003) | Y | N | N | N | N | N | N | N | N | N | Y | 2/11 Low |

Key: Y – Yes; N – No; U – Unclear; n/a – not applicable

1. Is the review question clearly and explicitly stated?
2. Were the inclusion criteria appropriate for the review question?
3. Was the search strategy appropriate?
4. Were the sources and resources used to search for studies adequate?
5. Were the criteria for appraising studies appropriate?
6. Was critical appraisal conducted by two or more reviewers independently?
7. Were there methods to minimize errors in data extraction?
8. Were the methods used to combine studies appropriate?
9. Was the likelihood of publication bias assessed?
10. Were recommendations for policy and/or practice supported by the reported data?
11. Were the specific directives for new research appropriate?

**NOTE:** Cut off scores are defined in this review as; 10-11 - high quality, 5 to 9 – moderate quality, 0 to 4 – low quality.

##### Table 6.2 JBI critical appraisal scores for qualitative studies (Joanna Briggs Institute, 2017b)

| **Study** | **JBI appraisal items** | | | | | | | | | | **Score** |
| --- | --- | --- | --- | --- | --- | --- | --- | --- | --- | --- | --- |
|  | **Q1** | **Q2** | **Q3** | **Q4** | **Q5** | **Q6** | **Q7** | **Q8** | **Q9** | **Q10** |  |
| Bryan et al. (2020) | Y | Y | Y | Y | Y | N | N | Y | N/A | Y | 7/10 Moderate |

Key: CT: Can’t tell; N: No; Y: Yes

Q1: Is there congruity between the stated philosophical perspective and the research methodology?

Q2: Is there congruity between the research methodology and the research question or objectives?

Q3: Is there congruity between the research methodology and the methods used to collect data?

Q4: Is there congruity between the research methodology and the representation and analysis of data?

Q5: Is there congruity between the research methodology and the interpretation of results?

Q6: Is there a statement locating the researcher culturally or theoretically?

Q7: Is the influence of the researcher on the research, and vice- versa, addressed?

Q8: Are participants, and their voices, adequately represented?

Q9: Is the research ethical according to current criteria or, for recent studies, and is there evidence of ethical approval by an appropriate body?

Q10: Do the conclusions drawn in the research report flow from the analysis, or interpretation, of the data?

**NOTE:** Cut off scores are defined in this review as; 9-10 high quality, 5 to 8 – moderate quality, 0 to 4 – low quality.

##### Table 6.3 JBI critical appraisal checklist for cross-sectional studies (Joanna Briggs Institute, 2017a).

| **Study** | **JBI Appraisal items** | | | | | | | | | | | **Score** |
| --- | --- | --- | --- | --- | --- | --- | --- | --- | --- | --- | --- | --- |
|  | **1** | **2** | **3** | **4** | **5** | **6** | **7** | **8** | **9** | **10** | **11** |  |
| Reacher et al. (2004) | Y | Y | Y | Y | Y | Y | Y | N/A | N/A | N/A | Y | 8/11 Moderate |

Key: Y: Yes; N: No; U: Unclear; n/a: not applicable

1. Were the two groups similar and recruited from the same population?
2. Were the exposures measured similarly to assign people to both exposed and unexposed groups?
3. Was the exposure measured in a valid and reliable way?
4. Were confounding factors identified?
5. Were strategies to deal with confounding factors stated?
6. Were the groups/participants free of the outcome at the start of the study (or at the moment of exposure)?
7. Were the outcomes measured in a valid and reliable way?
8. Was the follow up time reported and sufficient to be long enough for outcomes to occur?
9. Was follow up complete, and if not, were the reasons to loss to follow up described and explored?
10. Were strategies to address incomplete follow up utilized?
11. Was appropriate statistical analysis used?

**NOTE:** Cut off scores are defined in this review as; 10-11 - high quality, 5 to 9 – moderate quality, 0 to 4 – low quality.

##### Table 6.4 JBI critical appraisal checklist for descriptive surveys (Joanna Briggs Institute, 2017a)

| **Study** | **JBI Appraisal items** | | | | | | | | **Score** |
| --- | --- | --- | --- | --- | --- | --- | --- | --- | --- |
|  | **1** | **2** | **3** | **4** | **5** | **6** | **7** | **8** |  |
| Fewtrell et al. (2011) | Y | Y | Y | Y | Y | Y | Y | Y | 8/8 High |
| Findlater et al. (2023) | Y | Y | Y | N | Y | Y | Y | Y | 7/8 High |
| French et a (2019) | Y | Y | Y | Y | Y | Y | Y | Y | 8/8 High |
| Graham et al. (2019) | Y | Y | Y | Y | Y | Y | Y | Y | 8/8 High |
| Lamond et al. (2015) | Y | Y | Y | Y | Y | Y | Y | Y | 8/8 High |
| Mason et al. (2010) | Y | Y | Y | Y | Y | Y | Y | Y | 8/8 High |
| Munro et al. (2017) | Y | Y | Y | Y | Y | Y | Y | Y | 8/8 High |
| Mulchandani et al. (2020) | Y | Y | Y | Y | Y | Y | Y | Y | 8/8 High |
| Paranjothy et al. (2011) | Y | Y | Y | Y | Y | Y | Y | Y | 8/8 High |
| Robin et al. (2020) | Y | Y | Y | Y | Y | Y | Y | Y | 8/8 High |
| Waite et al. (2017) | Y | Y | Y | Y | Y | Y | Y | Y | 8/8 High |

Key: Y – Yes; N – No; U – Unclear; n/a – not applicable

1. Were the criteria for inclusion in the sample clearly defined?
2. Were the study subjects and the setting described in detail?
3. Was the exposure measured in a valid and reliable way?
4. Were objective, standard criteria used for measurement of the condition?
5. Were confounding factors identified?
6. Were strategies to deal with confounding factors stated?
7. Were the outcomes measured in a valid and reliable way?
8. Was appropriate statistical analysis used?

**NOTE:** Cut off scores are defined in this review as; 7 to 8 - high quality, 5 to 7 – moderate quality, 0 to 4 – low quality.

##### Table 6.5 JBI critical appraisal scores for qualitative studies (Joanna Briggs Institute, 2017b)

| **Study** | **JBI appraisal items** | | | | | | | | | | **Score** |
| --- | --- | --- | --- | --- | --- | --- | --- | --- | --- | --- | --- |
|  | **Q1** | **Q2** | **Q3** | **Q4** | **Q5** | **Q6** | **Q7** | **Q8** | **Q9** | **Q10** |  |
| Carroll et al. (2010) | Y | Y | Y | Y | Y | N | N | Y | Y | Y | 8/10  Moderate |
| Fothergill et al. (2021) | Y | Y | Y | Y | Y | N | N | Y | Y | Y | 8/10  Moderate |
| Medd et al. (2015) | Y | Y | Y | Y | Y | N | N | Y | N | Y | 7/10  Moderate |
| Mehring et al. (2023) | Y | Y | Y | Y | Y | Y | Y | Y | Y | Y | 10/10  High |
| Tapsell et al. (2002) | Y | Y | Y | Y | Y | N | N | N | N | Y | 6/10  Moderate |

Key: CT: Can’t tell; N: No; Y: Yes

Q1: Is there congruity between the stated philosophical perspective and the research methodology?

Q2: Is there congruity between the research methodology and the research question or objectives?

Q3: Is there congruity between the research methodology and the methods used to collect data?

Q4: Is there congruity between the research methodology and the representation and analysis of data?

Q5: Is there congruity between the research methodology and the interpretation of results?

Q6: Is there a statement locating the researcher culturally or theoretically?

Q7: Is the influence of the researcher on the research, and vice- versa, addressed?

Q8: Are participants, and their voices, adequately represented?

Q9: Is the research ethical according to current criteria or, for recent studies, and is there evidence of ethical approval by an appropriate body?

Q10: Do the conclusions drawn in the research report flow from the analysis, or interpretation, of the data?

**NOTE:** Cut off scores are defined in this review as; 9-10 high quality, 5 to 8 – moderate quality, 0 to 4 – low quality.

##### Table 6.6 JBI critical appraisal scores for quasi-experimental studies

| **Study** | **JBI Appraisal items** | | | | | | | | | **Score** |
| --- | --- | --- | --- | --- | --- | --- | --- | --- | --- | --- |
|  | **Q1** | **Q2** | **Q3** | **Q4** | **Q5** | **Q6** | **Q7** | **Q8** | **Q9** |  |
| Fewtrell and Kay (2008)  (Urban case study) | Y | Y | Y | N | N/A | N/A | Y | Y | Y | 6/9 Moderate |
| Milojevic et al. (2011) | Y | Y | Y | N | N | N/A | Y | Y | Y | 6/9 Moderate |
| Milojevic et al. (2017) | Y | Y | Y | N/A | N/A | N/A | Y | Y | Y | 6/9 Moderate |

Key: N: No; Y: Yes, U: Unclear

Q1: Is it clear in the study what is the ‘cause’ and what is the ‘effect’ (i.e. there is no confusion about which variable comes first)?

Q2: Were the participants included in any comparisons similar?

Q3: Were the participants included in any comparisons receiving similar treatment/care, other than the exposure or intervention of interest?

Q4: Was there a control group?

Q5: Were there multiple measurements of the outcome both pre and post the intervention/exposure?

Q6: Was follow up complete and if not, were differences between groups in terms of their follow up adequately described and analyzed?

Q7: Were the outcomes of participants included in any comparisons measured in the same way?

Q8: Were outcomes measured in a reliable way?

Q9: Was appropriate statistical analysis used?

**NOTE:** Cut off scores are defined in this review as; 8-9 - high quality, 5 to 7 – moderate quality, 0 to 4 – low quality.

#### Extreme heat studies: Quality appraisal

The Joanna Briggs quality appraisal tools for analytical cross-sectional studies and review studies were used for the following quality appraisals (The Joanna Briggs Institute, 2017a, 2017b). See Tables below.

##### Table 6.7 JBI critical appraisal checklist for systematic reviews and research synthesis (Joanna Briggs Institute, 2017a)

| **Study** | **JBI Appraisal items** | | | | | | | | | | | **Score** |
| --- | --- | --- | --- | --- | --- | --- | --- | --- | --- | --- | --- | --- |
|  | **1** | **2** | **3** | **4** | **5** | **6** | **7** | **8** | **9** | **10** | **11** |  |
| Arbuthnott et al. (2017) | Y | Y | Y | Y | N | N | N | Y | N | Y | Y | 7/11 Moderate |
| Cruz et al. (2019) | Y | Y | Y | Y | Y | N | N | Y | N | Y | Y | 8/11 Moderate |
| Curtis et al. (2017) | Y | N | N | N | N | N | N | Y | N | Y | Y | 4/11 Low |
| Finlay et al. (2012) | Y | Y | Y | Y | N | N | N | Y | Y | Y | Y | 8/11  Moderate |

Key: Y – Yes; N – No; U – Unclear; n/a – not applicable

12. Is the review question clearly and explicitly stated?
13. Were the inclusion criteria appropriate for the review question?
14. Was the search strategy appropriate?
15. Were the sources and resources used to search for studies adequate?
16. Were the criteria for appraising studies appropriate?
17. Was critical appraisal conducted by two or more reviewers independently?
18. Were there methods to minimize errors in data extraction?
19. Were the methods used to combine studies appropriate?
20. Was the likelihood of publication bias assessed?
21. Were recommendations for policy and/or practice supported by the reported data?
22. Were the specific directives for new research appropriate?

**NOTE:** Cut off scores are defined in this review as; 10-11 - high quality, 5 to 9 – moderate quality, 0 to 4 – low quality.

##### Table 6.8 JBI critical appraisal checklist for analytical cross-sectional studies (The Joanna Briggs Institute, 2017a)

| **Study** | **JBI Appraisal items** | | | | | | | | **Score** |
| --- | --- | --- | --- | --- | --- | --- | --- | --- | --- |
|  | **1** | **2** | **3** | **4** | **5** | **6** | **7** | **8** |  |
| Alahmad et al. (2023) | Y | Y | Y | Y | Y | Y | Y | Y | 8/8 High |
| Berger et al. (XXXX) | Y | Y | Y | Y | Y | Y | Y | Y | 8/8 High |
| Green et al. (2016) | Y | Y | Y | Y | Y | Y | Y | Y | 8/8 High |
| Hajat et al. (2002) | Y | Y | Y | Y | Y | Y | Y | Y | 8/8 High |
| Heaviside et al. (2016) | Y | Y | Y | Y | Y | Y | Y | Y | 8/8 High |
| Johnson et al. (2005) | Y | Y | Y | Y | N | N | Y | N | 5/8 Moderate |
| Kovats et al. (2004) | Y | Y | Y | Y | Y | Y | Y | Y | 8/8 High |
| Leonardi et al. (XXXX) | Y | Y | Y | Y | Y | Y | Y | Y | 8/8 High |
| Oven et al. (2012) | N | Y | Y | Y | N | N | N | N | 8/8 High |
| Page et al. (2007) | Y | Y | Y | Y | Y | Y | Y | Y | 8/8 High |
| Page et al. (2012) | Y | Y | Y | Y | Y | Y | Y | Y | 8/8 High |
| Rendell et al. (2020) | Y | Y | Y | Y | Y | Y | Y | Y | 8/8 High |
| Rooney et al. (1998) | Y | Y | Y | Y | Y | Y | Y | Y | 8/8 High |
| Rizmie et al. (2022) | Y | Y | Y | Y | Y | Y | Y | Y | 8/8 High |
| Sahani et al. (2022) | Y | Y | Y | Y | Y | Y | Y | Y | 8/8 High |
| Smith et al. (2016) - (Estimating the burden) | Y | Y | Y | Y | Y | Y | Y | Y | 8/8 High |
| Smith et al. (2016) - (The impact of heatwaves on healthcare usage) | Y | Y | Y | Y | Y | Y | Y | Y | 8/8 High |
| Thompson et al. (2022) | Y | Y | Y | Y | Y | Y | Y | Y | 8/8 High |
| Zhang et al. (2023) | Y | Y | Y | Y | Y | Y | Y | Y | 8/8 High |
| Wan et al. (2022) | Y | Y | Y | Y | Y | Y | Y | Y | 8/8 High |

Key: Y – Yes; N – No; U – Unclear; n/a – not applicable

1. Were the criteria for inclusion in the sample clearly defined?
2. Were the study subjects and the setting described in detail?
3. Was the exposure measured in a valid and reliable way?
4. Were objective, standard criteria used for measurement of the condition?
5. Were confounding factors identified?
6. Were strategies to deal with confounding factors stated?
7. Were the outcomes measured in a valid and reliable way?
8. Was appropriate statistical analysis used?

**NOTE:** Cut off scores are defined in this review as; 8-9 - high quality, 5 to 7 – moderate quality, 0 to 4 – low quality.

#### Extreme wind study

For the extreme wind included study, the quality appraisal tool used was the JBI critical appraisal scores for systematic reviews and research synthesis (The Joanna Briggs Institute, 2017c).

##### Table 6.9 JBI critical appraisal checklist for analytical cross-sectional studies (The Joanna Briggs Institute, 2017a)

| **Study** | **JBI appraisal items** | | | | | | | | | | | **Score** |
| --- | --- | --- | --- | --- | --- | --- | --- | --- | --- | --- | --- | --- |
|  | **Q1** | **Q2** | **Q3** | **Q4** | **Q5** | **Q6** | **Q7** | **Q8** | **Q9** | **Q10** | **Q11** |  |
| Goldman et al. (2014) | Y | Y | Y | Y | Y | U | U | Y | U | Y | Y | 8/11  Moderate |

Key: Y – Yes; N – No; U – Unclear; n/a – not applicable

Q1: Is the review question clearly and explicitly stated?

Q2: Were the inclusion criteria appropriate for the review question?

Q3: Was the search strategy appropriate?

Q4: Were the sources and resources used to search for studies adequate?

Q5: Were the criteria for appraising studies appropriate?

Q6: Was critical appraisal conducted by two or more reviewers independently?

Q7: Were there methods to minimize errors in data extraction?

Q8: Were the methods used to combine studies appropriate?

Q9: Was the likelihood of publication bias assessed?

Q10: Were recommendations for policy and/or practice supported by the reported data?

Q11: Were the specific directives for new research appropriate?

**NOTE:** Cut off scores are defined in this review as; 10-11 - high quality, 5 to 9 – moderate quality, 0 to 4 – low quality.

**END**
